# Noninvasive ventilation strategies for patients with severe or critical COVID-19: A rapid review of clinical outcomes

**DOI:** 10.1101/2022.05.25.22275586

**Authors:** Shannon E. Kelly, George A. Wells

## Abstract

**Objectives:** To examine whether high flow nasal oxygen (HFNO), continuous positive airway pressure (CPAP), or noninvasive ventilation (NIV) strategies impact mortality, the need for invasive mechanical ventilation (IMV), or hospital and intensive care unit (ICU) length of stay compared to standard oxygen therapy (SOT) or each other in patients with severe or critical COVID-19 with acute hypoxemic respiratory failure.

**Methods:** A rapid review of randomized controlled trials (RCTs) identified through published systematic and rapid reviews supplemented with a search of bibliographic databases. RCTs were eligible if they compared HFNO, CPAP, or NIV to SOT or another ventilation strategy. Studies were screened, selected, and extracted by a single reviewer and checked by a second reviewer. We assessed risk of bias of included studies using the Cochrane ‘Risk of bias’ tool and used the grading of recommendations, assessment, development, and evaluation (GRADE) approach to judge the certainty of the evidence for mortality, need for IMV, and hospital and ICU length of stay. We sought RCT evidence for non-COVID-19 patients with acute hypoxemic respiratory failure and acute respiratory distress to inform additional comparisons and to supplement the available data for COVID-19.

**Results:** A total of 5 RCTs comparing ventilation strategies in patients with severe or critical COVID-19 were included. Patient and study characteristics were extracted and evidence and certainty of evidence assessments were completed for comparisons of HFNO and CPAP to standard oxygen therapy and NIV and CPAP to HFNO. An additional 22 RCTs of non-COVID-19 patients were also included and considered.

Results from meta-analysis suggest reductions in mortality and IMV with HFNO (*RR mortality 0*.*87 (0*.*66-1*.*13), IMV 0*.*89 (0*.*77-1*.*03); low quality evidence*) or CPAP (*RR mortality 0*.*87 (0*.*64-1*.*18) low quality evidence, IMV 0*.*81 (0*.*67-0*.*98) moderate quality evidence)* compared to SOT. Helmet NIV may reduce IMV (*RR 0*.*69 (0*.*43-1*.*09))* and CPAP may reduce IMV *(RR 0*.*69 (0*.*43-1*.*09))* and hospital *(1*.*67 days fewer (5*.*43 fewer-2*.*09 more)* or ICU length of stay *(1*.*02 days fewer (3*.*97 fewer-1*.*93 more))* compared to HFNO *(low quality evidence)*.

**Conclusions:** This rapid systematic review highlights the available evidence to support the use of noninvasive ventilation strategies including high flow nasal oxygen, noninvasive ventiltaion (e.g., BiPAP), or CPAP in hospitalized patients with severe or critical COVID-19 and acute hypoxemic respiratory failure who do not need emergent intubation. Findings based on moderate to very low certainty evidence suggest that noninvasive ventilation may be considered as an alternative to standard oxygen therapy to reduce hypoxemia and dyspnea. Additional high quality RCTs are warranted to reduce uncertainty and to fill in important knowledge gaps.

## Introduction

Severe acute respiratory syndrome coronavirus 2 (SARS-CoV-2) emerged at the end of 2019 as a novel coronavirus, resulting in the current global pandemic of respiratory illness, Coronavirus Disease 2019 (COVID-19). Severe and critical COVID-19 involves acute hypoxemic respiratory failure requiring oxygen and ventilation therapies1. COVID-19 patients may deteriorate quickly and require hospitalization to prevent progressive hypoxemia, leading to acute respiratory failure^2^.

In addition to self or supported repositioning (i.e., prone positioning) and medication, support for patients with COVID-19 related hypoxemia includes oxygen therapy which often requires progressive increases in respiratory support. Clinical management of COVID-19 patients not responding well to standard oxygen may be considered for alternative ventilation support when hypoxemia is a concern. This ventilation may include high-flow nasal oxygen (HFNO), continuous positive airways pressure (CPAP), non-invasive positive pressure ventilation (NIV), invasive mechanical ventilation (IMV), or possibly a combination of more than one strategy^3^. Different interfaces, including oral, nasal or full face masks and helmets, may be used and are selected to minimize air leaks and adverse effects and maximize comfort and communication^4,5^. Care for patients with COVID-19 and acute respiratory failure requires intensive monitoring and management of blood oxygen. Based on experience early in the pandemic, we know that being intubated too early can result in poor outcomes for COVID-19 patients^6^. The case fatality rate for patients with COVID-19 admitted to the intensive care unit (ICU) and receiving IMV varies but is high^7^ and so avoiding progression to IMV is a common clinical goal in hospitalized patients with COVID-19.

Although there has been a redirection of attention in clinical research toward patients with COVID-19 since early 2020, there is an ongoing need to synthesize the research on non-invasive ventilation strategies for hospitalized patients with COVID-19 to inform clinical practice guidelines and care protocols. This rapid review aims to examine whether HFNO, CPAP, or NIV impact mortality, the need for IMV, or hospital and ICU length of stay (LOS) compared to standard oxygen therapy (SOT) or each other in patients with severe or critical COVID-19. In the absence of high certainty evidence for hospitalized patients with COVID-19 and acute hypoxemic respiratory failure (AHRF), we will consider the evidence for hospitalized patients with acute respiratory distress syndrome (ARDS) and AHRF.

## Methods

We conducted a rapid review of non-invasive ventilation strategies in patients with severe or critical COVID-19. Patient partners reviewed and provided feedback on the protocol registered with the National Collaborating Centre for Methods and Tools (NCCMT; Registration No. 428^8^).

The review team used a population, intervention, comparator, and outcomes (PICO) framework (**Table 1)**. We aimed to find, assess and synthesize all randomized controlled trials (RCTs) of severe or critical COVID-19 and acute hypoxemic respiratory failure (AHRF). Eligible studies were RCTs of patients hospitalized with severe or critical COVID-19 and AHRF not needing emergent intubation who used HFNO, CPAP or NIV (e.g., Bilevel Positive Airway Pressure; BiPAP). Comparisons of interest were standard oxygen therapy (SOT) or another intervention of interest. The primary outcomes are mortality, need for invasive mechanical ventilation (IMV), and hospital length of stay, with intensive care unit (ICU) length of stay and patient-reported outcomes (e.g. patient comfort, satisfaction with care) are secondary outcomes. Patients weaned off IMV or those who require respiratory support following IMV were not eligible. Reviewers also sought evidence in a related clinical population of individuals with ARDS and AHRF to provide indirect evidence for the use of noninvasive ventilation in COVID-19 patients (Methods and results, **Online Supplement 1**).

**Table 1.**
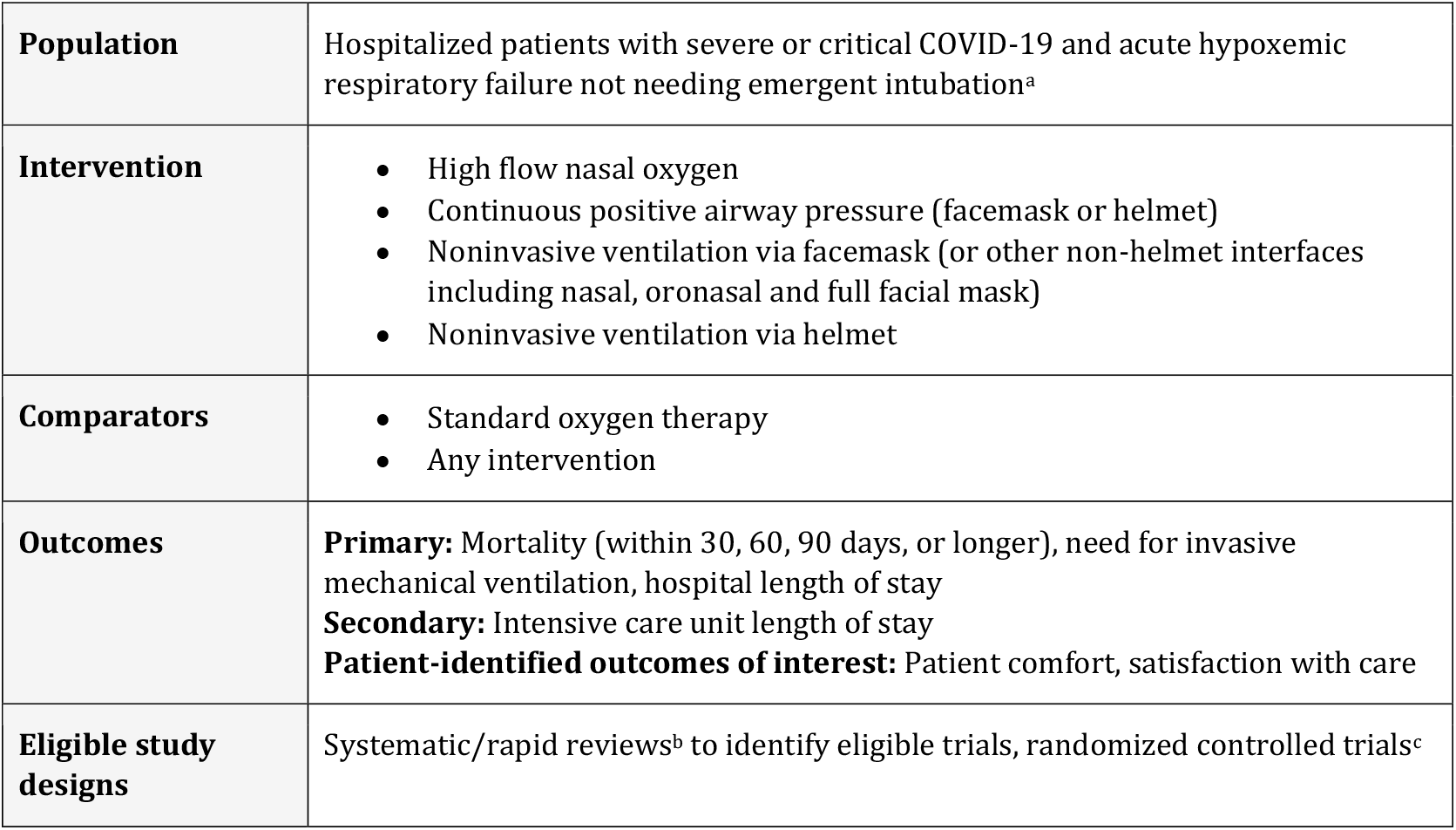
Population, intervention, comparator, outcomes (PICO) framework

|  |  |
| --- | --- |
| **Population** | Hospitalized patients with severe or critical COVID-19 and acute hypoxemic respiratory failure not needing emergent intubation<sup>a</sup> |
| **Intervention** | • High flow nasal oxygen<br>• Continuous positive airway pressure (facemask or helmet)<br>• Noninvasive ventilation via facemask (or other non-helmet interfaces including nasal, oronasal and full facial mask)<br>• Noninvasive ventilation via helmet |
| **Comparators** | • Standard oxygen therapy<br>• Any intervention |
| **Outcomes** | **Primary:** Mortality (within 30, 60, 90 days, or longer), need for invasive mechanical ventilation, hospital length of stay<br>**Secondary:** Intensive care unit length of stay<br>**Patient-identified outcomes of interest:** Patient comfort, satisfaction with care |
| **Eligible study designs** | Systematic/rapid reviews<sup>b</sup> to identify eligible trials, randomized controlled trials<sup>c</sup> |

## Search

### An experienced information scientist designed and executed the literature searches

The search first targeted systematic and rapid reviews (SR/RRs) of potentially eligible RCTs in two electronic COVID-19 databases (WHO COVID-19 Global literature on coronavirus disease database^9^, the Living OVerview of Evidence (L.OVE) platform)^10^ and the COVID-END inventory^11^ of best evidence syntheses for clinical management to rapidly identify eligible SR/RRs [May 2 to 3, 2021] (**Online supplement 2**). Reviewers checked the included study lists of the identified English language SR/RRs for potentially eligible RCTs and checked all ongoing RCT records from the SR/RRs for results (July 15, 2021).

A second search (WHO COVID-19 study register, Cochrane COVID-19 study register, clinicaltrials.gov; May 15, 2021) sought to identify RCTs published or registered since the last identified SR/RR search (01 July 2020). The information scientist did not execute the planned search of the International Clinical Trials Registry Platform (ICTRP)^12^ as the database was not accessible to our information scientist during this rapid review. We did not apply restrictions to publication status or language in the search strategy.

All ongoing studies identified were checked for results (June 15-21, 2021). We used monthly alerts to identify additional RCT records and performed a forward citation analysis on all included records using both SCOPUS and Google Scholar. Monitoring of search alerts and ongoing RCTs continued monthly until December 29, 2021.

### Study selection

All records were uploaded into EndNoteX20^13^ and then into Covidence^14^ for screening and selection. One reviewer screened all title and abstract records (SR/RRs and then RCTs) and confirmed the results with a second reviewer. One reviewer located and screened all potentially relevant full-text articles identified through the SR/RR and RCT searches. A second reviewer confirmed all included SR/RRs and RCTs and resolved disagreements through discussion. Reviewers used a PRISMA flow diagram to document the selection process in detail.

Included SR/RRs and RCTs directly compared two or more interventions or comparators in the PICO and reported at least one outcome of interest. We excluded studies reporting broad clinical course as it is not possible to link clinical outcomes and isolate the effect of the noninvasive ventilation strategy.

### Data extraction

For each included SR/RR, one reviewer extracted details of the citation, synthesis approach, PICO, search dates, and the number of included potentially relevant RCTs studies into structured summary tables (Microsoft Word).

For each included RCT, one reviewer retrieved all reported participant characteristics, study characteristics and outcome data from the SR/RR publication or extracted data de novo into structured outcome tables (Microsoft Excel). A second reviewer checked all extracted data.

### Risk of bias assessment

One reviewer appraised each included SR/RR using AMSTAR2^15^ and retrieved the risk of bias (ROB) assessments for the included RCTs where reported. Otherwise, a single reviewer assessed the ROB in trials with no retrieved assessment (Cochrane ROB tool^16^) or when the retrieved assessment did not have sufficient detail to inform the summary of findings table. ROB ratings were not used as exclusion criteria.

### Evidence tables and certainty of evidence assessment

One reviewer constructed GRADE evidence profiles of certainty for each primary and secondary outcome and each intervention comparison, separately using MAGICapp^17^. A second reviewer checked all tables, and they were vetted by content and methodological experts. The certainty of the evidence was determined by the risk of bias across studies, inconsistency, indirectness, and imprecision^18^. Reviewers did not consider incoherence, publication bias and other parameters. We used the most recent GRADE methodology to decide on the certainty of the body of evidence from RCTs, which recommends using the judgment of high certainty of evidence at baseline and downgrading due to risk of bias, imprecision, or indirectness of RCTs^18^. Minimal important differences and optimal information size (OIS; 1000 participants) were determined in advance through consult with content experts and used to inform judgements about precision. Downgrading 3 levels for precision was considered when the number of study participants for an outcome was less than 30% of the OIS^19^. When the certainty (or quality) of evidence is rated very low or extremely low, we are very unsure about the true effect, while a rating of low certainty/quality evidence means that the true effect could be markedly different from the estimated effect. Ratings of moderate certainty indicate that reviewers feel that the true effect is probably close to the estimated effect, and high certainty ratings signify a high level of confidence in the estimated effect^20^. Plain language statements are determined based on the ratings for the certainty of evidence and are presented with the ratings.

### Synthesis

Reviewers used Review Manager (Version 5.4; RevMan) to calculate the treatment effect for each primary and secondary outcome (risk ratios for dichotomous outcomes, mean difference for continuous outcomes). Reviewers undertook meta-analyses when 2 studies or comparisons reported the same outcome. A fixed effects model was used for all meta-analyses and the unit of analysis was individuals. Otherwise, reviewers completed a descriptive summary of evidence by comparison and outcome.

Treatment effects were calculated for the following comparisons: HFNO versus SOT, CPAP versus SOT, NIV versus SOT, CPAP versus HFNO, NIV versus HFNO, and NIV versus CPAP. The interface used for the non-invasive ventilation strategy was also compared if reported (e.g., helmet CPAP versus HFNO, facemask CPAP versus helmet CPAP).

When an RCT did not directly compare the study interventions (e.g., a 3-arm study where HFNO and NIV are each compared to SOT but not to each other), the relative effects were calculated using the pooled relative effects (risk ratio) for the interventions reported to calculate an indirect estimate for the relative effects of the comparison of interest^21^.

For RCTs reporting hospital or ICU LOS outcomes both as means (standard deviation) and medians (interquartile range; IQR) for any one outcome, meta-analysis was conducted in R using the Metamedian package^22,23^. LOS data were also checked for consideration of competing risk^24^.

The I^2^ statistic was used to measure heterogeneity among the included trials. Publication bias was not assessed or small studies effects because fewer than 10 studies were included. Subgroup or sensitivity analyses were not conducted, and investigators or study sponsors were not contacted to provide missing data.

## Results

A total of 342 title and abstract records and 41 full-text articles were screened for relevant SR/RRs. From these, we identified three SRs (reported in five published reports)^25-29^ and four RRs^30-33^ reporting non-invasive ventilation strategies in hospitalized patients with severe or critical COVID-19 and acute hypoxemic respiratory failure not requiring emergent intubation. Reviewers identified and included three RCTs^34-36^ in three records from the included study lists of these reviews and through related forward citation tracking. An additonal search for RCTs published between July 1, 2020 (the search date from the most recent included SR) and May 15, 2021 identified 1,385 title and abstract records (including records identified by alerts). Following screening, 24 full-text articles were reviewed and two more eligible RCTs were identified^37,38^. A total of five RCTs were included.

Three additional records reporting RCTs in-progress (NCT04326075, NCT04381923 and NCT04507802) and one terminated RCT^39^ were also located but were excluded as no posted or published results were available within our search or alert time-frame.

The screening and selection process is reported in a PRISMA 2020 flow diagram modified to capture the search for SR/RRs and RCTs (**Figure 1**)^40^. The included SR/RRs and their AMSTAR2 assessments are summarized in **Online Supplement 3**.

**Figure 1.**
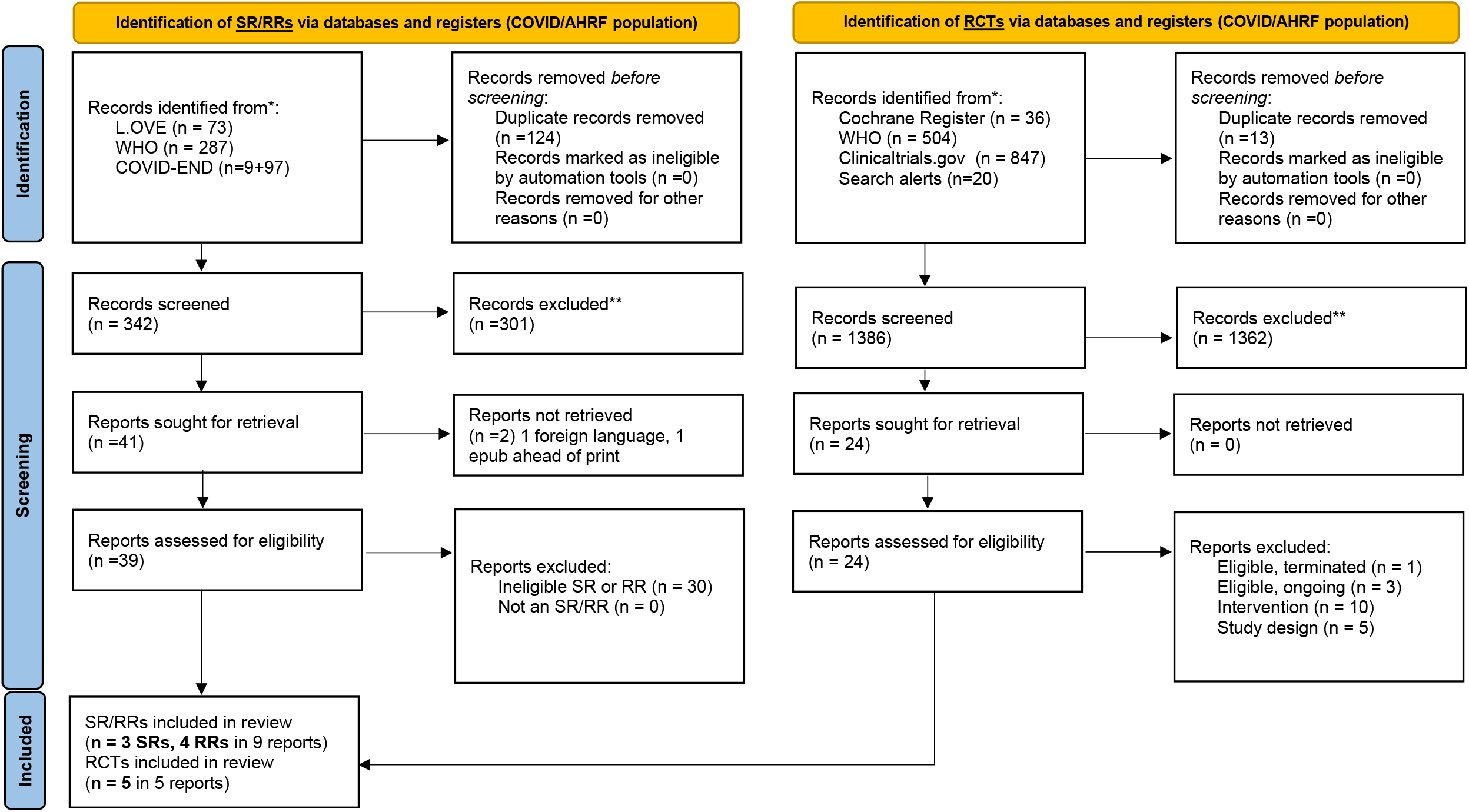
Modified PRISMA 2020 flow diagram for identification of RCTs of COVID-19 and AHRF population^40^.

### Study and participant characteristics

A summary of the five included RCTs^34-38^ is provided in **Table 2**. Additional details for the study and participant characteristics are reported in **Online Supplement 4**. Four of the RCTs compared HFNO to standard oxygen therapy^34-36,38^, one 3-arm RCT compared CPAP to standard oxygen therapy^35^, one 2-arm RCT compared helmet NIV to HFNO^37^ and one three-arm RCT compared CPAP and HFNO^35^ to SOT, but not directly to each other. For the comparison of CPAP to HFNO, an indirect treatment comparison estimate was made. Not all RCTs reported all outcomes.

**Table 2.** Characteristics of included RCTs

| Study/Design | Population | Country/Setting | Interventions | Outcomes reported |
| --- | --- | --- | --- | --- |
| **Li et al. 2020**<sup>34</sup><br>two-arm, parallel RCT<br>N=72 | Patients with severe coronavirus pneumonia complicated with acute respiratory failure | China, isolation ward of a single centre | HFNO [n=37]<br>Standard oxygen therapy [n=35] | Mechanical ventilation at 12 h<br>No patient-reported outcomes |
| **Grieco et al. 2021**<sup>37</sup><br>HENIVOT<br>two-arm, parallel RCT<br>N=109 | Patients admitted to the intensive care unit with COVID-19-induced moderate to severe hypoxemic respiratory failure | Italy, ICUs in four centres | Helmet NIV [n=55]<br>HFNO [n=54] | Intubation, 28 d<br>Hospital LOS<br>ICU LOS<br>Patient-reported: Device-related discomfort |
| **Perkins et al. 2021**<sup>35</sup><br>RECOVERY-RS<br>three-arm, adaptive RCT<br>N=1272 | Hospitalized adults with acute respiratory failure due to COVID-19 were deemed suitable for tracheal intubation if treatment escalation was required | United Kingdom, 75 hospitals | CPAP [n=380]<br>HFNO [n=417]<br>Standard oxygen therapy [n=475]<br>(primary comparisons were CPAP to standard oxygen and HFNO to standard oxygen) | Mortality, 30 d<br>Intubation, 30 d<br>Tracheal intubation during the study period<br>Critical care (ICU) LOS<br>Hospital LOS<br>No patient-reported outcomes |
| **Teng et al. 2021**<sup>36</sup><br>two-arm, parallel RCT | Patients diagnosed with severe COVID-19. | China, single centre | HFNO [n=12]<br>Standard oxygen therapy [n=10] | Mortality (indirect)<br>Hospital LOS |
| N= 22 |  |  |  | ICU LOS<br>No patient-reported outcomes |
| **Ospina-Tascón et al. 2021**<sup>38</sup><br>Two-arm, open-label parallel RCT<br>N=199 | Adult patients admitted to the emergency department, general ward, or intensive care unit with acute respiratory failure and COVID-19 | Colombia, three centres | HFNO [n=99]<br>Standard oxygen therapy [n=100] | Mortality, 28 d<br>Intubation, 28 d<br>Hospital LOS<br>ICU LOS<br>No patient-reported outcomes |

d=days; h=hours; HFNO=high flow nasal oxygen; ICU=intensive care unit; LOS=length of stay; RCT=randomized controlled trial; QoL=quality of life.

Li et al. (2020) is a two-arm, parallel RCT conducted in 72 patients with severe COVID-19 pneumonia and acute respiratory failure in the Huanggang Hospital in China, where participants were randomized to either HFNO (n=37) or SOT (n=35)^34^. The Li et al. was published in simplified Chinese and all eligibility, data extraction and risk of bias assessments were completed by a fluent reviewer. The study reported one outcome: mechanical ventilation outcomes at 12 hours.

Perkins et al. (2021; RECOVERY-RS) is a three-arm, open-label, adaptive RCT conducted in 75 centres in the United Kingdom. A total of 1,272 individuals were enrolled and randomized to CPAP, HFNO, or SOT. The primary comparisons in the study are CPAP to SOT and HFNO to SOT. All primary and secondary outcomes were reported. Results from the pre-print of this RCT, which was published in August 2021, were originally included^35^ and have been checked for accuracy against the published and peer-reviewed RCT which was published in January 2022^41^.

Teng et al. (2021) is a two-arm, parallel RCT comparing HFNO to SOT in 22 patients diagnosed with severe COVID-19 at a single centre in China^36^. The trial reported LOS outcomes and that all participants were cured and discharged which was inferred as a zero mortality outcome in both study arms.

The HENIVOT trial by Grieco et al. (2021) randomized ICU patients in 4 centres in Italy with COVID-19 induced moderate to severe hypoxemic respiratory failure to NIV with a helmet interface or HFNO^37^. This open-label, multicenter, RCT assessed whether early treatment with helmet NIV decreased use of IMV in the 28 days after randomization compared to HFNO. This study reported all primary and secondary outcomes of interest and was the only RCT to report a patient-important outcome (device-related discomfort).

The two-arm parallel open label RCT by Ospina-Tascón et al. (HiFLo COVID; 2021) compared HFNO to SOT in 199 adult patients with acute respiratory failure and COVID-19^38^. Participants were adults admitted to the emergency department, general ward, or intensive care unit of 3 centres in Columbia. This study reported all primary and secondary outcomes of interest.

### Risk of Bias

There was limited information regarding the assessed ROB of the individual RCTs to carry forward from the SR/RRs, and therefore de novo ROB assessments were individually completed (**Online Supplement 5)**. Mortality and IMV outcomes were considered in the assessment of blinding at the participant and personnel level, and IMV was specifically considered when blinding of outcome assessment was rated. Participants and personnel were not blinded in any of the included RCTs (and blinding would have been difficult or impossible due to the nature of the interventions). Varying criteria may have been used within or across studies to initiate IMV. Intubation outcomes in the Grieco et al. RCT were independently adjudicated by external experts, but no adjudication of the intubation outcomes was reported in the other RCTs. Crossover or progression to other interventions or study arms and cointerventions could not be fulsomely assessed in the included RCTs, and therefore the risk of bias is unclear. Intention to treat data are extracted where available, but the effects in some patients may be attributable to multiple therapies.

### Evidence Tables

Evidence and certainty of evidence assessments for all comparisons and outcomes are summarized in **Table 3** (HFNO v. SOT, CPAP v. SOT, NIV v. HFNO, CPAP v. HFNO). Detailed individual outcome tables for all primary (mortality, IMV, hospital LOS) and secondary (ICU LOS) outcomes are provided in **Online Supplement 6**.

**Table 3.** Evidence and certainty of evidence.

|  |  | Quality assessment |  |  |  | Effect |  |  |  |  |  |  |
| --- | --- | --- | --- | --- | --- | --- | --- | --- | --- | --- | --- | --- |
| Outcome | No. of participants (studies) | Risk of bias | Inconsistency | Indirectness | Imprecision | Relative (95% CI) | Absolute Difference | Absolute effect intervention | Absolute effect intervention | Certainty (Quality) of the evidence | Importance | Plain language summary |
| HFNO vs. SOT, 4 RCTs, 1,053 participants<sup>34-36,38</sup> |  |  |  |  |  |  |  | SOT | HFNO |  |  |  |
| Mortality | 1,006 (3 RCTs) | No concerns | No concerns | No concerns | Very serious | RR 0.87 (0.66 - 1.13) | 24 fewer per 1000 (64 fewer - 24 more) | 188 per 1000 | 164 per 1000 | Low | Critical | HFNO may decrease mortality |
| IMV | 1,053 (3 RCTs) | No concerns | No concerns | No concerns | Very serious | RR 0.89 (0.77 - 1.03) | 46 fewer per 1000 (96 fewer - 13 more) | 417 per 1000 | 371 per 1000 | Low | Critical | HFNO may decrease IMV |
| Hospital LOS | 1,003 (3 RCTs) | No concerns | No concerns | No concerns | Serious | -- | 1.08 fewer days (2.48 fewer – 0.35 more) | 16.85 days | 16.85 days | Moderate | Critical | HFNO probably decreases hospital LOS |
| ICU LOS | 1,003 (3 RCTs) | No concerns | No concerns | No concerns | Serious | -- | 0.77 fewer days (1.45 fewer – 0.08 fewer) | 5.83 days | 4.65 days | Moderate | Important | HFNO probably has little or no difference in ICU LOS |
| CPAP vs. SOT, 1 RCT, 1,273 participants<sup>35</sup> |  |  |  |  |  |  |  | SOT | CPAP |  |  |  |
| Mortality | 737 (1 RCT) | No concerns | No concerns | No concerns | Very serious | RR 0.87 (0.64 - 1.18) | 25 fewer per 1000 (69 fewer - 35 more) | 192 per 1000 | 167 per 1000 | Low | Critical | CPAP may decrease mortality |
| IMV | 733 (1 RCT) | No concerns | No concerns | No concerns | Serious | RR 0.81 (0.67 - 0.98) | 78 fewer per 1000 (69 fewer - 8 fewer) | 413 per 1000 | 335 per 1000 | Moderate | Critical | CPAP probably decreases IMV |
| Hospital LOS | 737 (1 RCT) | No concerns | No concerns | No concerns | Very serious | -- | 0.96 fewer (3.59 fewer – 1.67 more) | 17.3 days | 16.4 days | Low | Critical | CPAP may decrease hospital LOS |
| ICU LOS | 737 (1 RCT) | No concerns | No concerns | No concerns | Very serious | -- | 0.08 fewer (2.23 fewer – 2.07 more) | 9.6 days | 9.5 days | Low | Important | CPAP may have little or no difference on ICU LOS |
| Outcome | No. of participants (studies) | Risk of bias | Inconsistency | Indirectness | Imprecision | Relative (95% CI) | Absolute Difference | Absolute effect intervention | Absolute effect intervention | Certainty (Quality) of the evidence | Importance | Plain language summary |
| **Helmet NIV vs. HFNO, 1 RCT, 110 participants<sup>37</sup>** |  |  |  |  |  |  |  | **HFNO** | **Helmet NIV** |  |  |  |
| Mortality | 110 (1 RCT) | No concerns | No concerns | No concerns | Extremely serious | RR 1.1 (0.55 - 2.2) | 24 more per 1000 (106 fewer - 283 more) | 236 per 1000 | 260 per 1000 | Very low | Critical | We are very uncertain of the impact of helmet NIV on mortality |
| IMV | 110 (1 RCT) | No concerns | No concerns | No concerns | Very serious | RR 0.54 (0.32 - 0.89) | 234 fewer per 1000 (346 fewer - 56 fewer) | 509 per 1000 | 275 per 1000 | Low | Critical | Helmet NIV may decrease IMV |
| Hospital LOS | 110 (1 RCT) | No concerns | No concerns | No concerns | Extremely serious | -- | 1 fewer (9.2 fewer - 7.2 more) | 22 days | 21 days | Very low | Critical | We are very uncertain of the impact of Helmet NIV on hospital LOS |
| ICU LOS | 110 (1 RCT) | No concerns | No concerns | No concerns | Extremely serious | -- | 1 fewer (6.2 fewer - 7.3 more) | 10 days | 9 days | Very low | Important | We are very uncertain of the impact of Helmet NIV on ICU LOS |
| Device-related discomfort | 110 (1 RCT) | Serious | No concerns | No concerns | Serious | -- | 1.9 higher (1.4 higher -2.5 higher) | 1.8 VAS points | 3.7 VAS points | Low | Patient-important | Helmet NIV may increase device-related discomfort |
| **CPAP vs. HFNO, 1 RCT, 1,273 participants<sup>35</sup>** |  |  |  |  |  |  |  | **CPAP** | **HFNO** |  |  |  |
| Mortality | 793 (1 RCT) | No concerns | No concerns | No concerns | Extremely serious | RR : 0.95 (0.52 - 1.71) | 9 fewer per 1000 (90 fewer - 133 more) | 188 per 1000 | 179 per 1000 | Very low | Critical | We are very uncertain of the impact of CPAP on mortality |
| IMV | 791 (1 RCT) | No concerns | No concerns | No concerns | Very serious | RR 0.69 (0.43 - 1.09) | 127 fewer per 1000 (234 fewer - 37 more) | 411 per 1000 | 284 per 1000 | Low | Critical | CPAP may decrease IMV |
| Hospital LOS | 791 (1 RCT) | No concerns | No concerns | No concerns | Very serious | -- | 1.67 fewer (5.43 fewer - 2.09 more) | 18.3 days | 16.4 days | Low | Critical | CPAP may decrease hospital LOS |
| Outcome | No. of participants (studies) | Risk of bias | Inconsistency | Indirectness | Imprecision | Relative (95% CI) | Absolute Difference | Absolute effect intervention | Absolute effect intervention | Certainty (Quality) of the evidence | Importance | Plain language summary |
| ICU LOS | 791 (1 RCT) | No concerns | No concerns | No concerns | Very serious | -- | 1.02 fewer (3.97 fewer - 1.93 more) | 10.5 days | 9.5 days | Low | Important | CPAP may decrease ICU LOS |
| *CI = confidence interval; CPAP = continuous positive airway pressure; HFNO = high flow nasal oxygen; ICU = intensive care unit; IMV = invasive mechanical ventilation; LOS = length of stay; NIV = noninvasive ventilation; No. = number; RCT = randomized controlled trial; RR = relative risk; SOT = standard oxygen therapy; VAS = visual analogue scale* |  |  |  |  |  |  |  |  |  |  |  |  |

### HFNO versus SOT

Four published RCTs compared HFNO to SOT (1053 participants) in patients with severe COVID-19. Based on low quality evidence from 3 RCTs, HFNO may decrease mortality (relative risk (RR) 0.87, 95% confidence interval (CI) 0.66 - 1.13) and IMV (RR 0.89, 95% CI 0.77 - 1.03) compared to SOT. HFNO probably decreases hospital LOS (1.08 fewer days, 95% CI 2.48 fewer – 0.35 more)^34-36,38^. HFNO probably has little or no difference in ICU LOS (moderate quality evidence). None of the trials included children or pregnant women, and none reported any patient-important outcomes (e.g., comfort, satisfaction).

Evidence from five additional RCTs conducted in non-COVID patients with ARDS and AHRF (1425 participants) is generally consistent, although there is less certainty over potential differences in mortality^42-47^.

### CPAP versus SOT

One published RCT of 742 participants compared CPAP to SOT^35^. Based on moderate quality evidence, CPAP probably decreases IMV compared to SOT (RR 0.81, 95% CI 0.67 - 0.98). There is low quality evidence that CPAP may decrease mortality (RR 0.87, 95% CI 0.64 - 1.18) and hospital LOS (0.96 fewer days; 95% CI 3.59 fewer – 1.67 more) and may have little or no difference in ICU LOS (0.08 fewer days, 95% CI 2.23 fewer – 2.07 more; low quality evidence). The study did not report any patient-important outcomes and did not include pregnant women or children.

Four RCTs of non-COVID ARDS and AHRF patients compared CPAP to SOT^48-51^. Three RCTs compared helmet CPAP interfaces to SOT (168 participants) and one RCT compared a face mask CPAP interface to SOT (123 patients)^48-51^. Evidence was very uncertain for all outcomes except hospital LOS where helmet CPAP may have little or no difference compared to SOT (0.5 more days, 95% CI 3.75 fewer - 4.75 more).

### NIV versus SOT

#### No RCTs compared NIV to SOT in patients with COVID-19

In 11 RCTs (1254 participants) of non-COVID ARDS and AHRF patients, NIV with a face mask interface probably decreases mortality (RR 0.83, 95% CI 0.71 – 0.96) and IMV (RR 0.74, 95% CI 0.64 - 0.86) compared to SOT (moderate quality evidence) and may decrease hospital (2.02 days fewer, 95% CI 4.39 fewer – 0.35 more) and ICU LOS (1.61 fewer days, 95% CI 3.21 fewer – 0.03 more)(low quality evidence)^43,45,52-61^. There was no evidence for children or pregnant women, or any non-facemask NIV interfaces.

### NIV versus HFNO

One small RCT compared NIV with a helmet interface to HFNO for patients with severe COVID-19 (110 participants)^37^. Helmet NIV may decrease the need for IMV (RR 0.54, 95% CI 0.32 – 0.89) but may increase device-related discomfort (low quality evidence). The impact of helmet NIV on mortality and hospital or ICU LOS compared to HFNO in COVID-19 patients is very uncertain.

Findings for mortality and IMV were very uncertain in three RCTs of non-COVID ARDS and AHRF patients (316 participants) comparing face mask NIV to HFNO, although there may be little or no difference in ICU LOS (1 RCT, 216 participants; low quality evidence)^45,62,63^. No hospital LOS data were reported.

### CPAP versus HFNO

One RCT^35,41^ enrolled 1273 participants into SOT, HFNO and CPAP arms but did not directly compare CPAP to HFNO in the trial analyses. As such, the meta-analysis for the comparison of CPAP vs. HFNO was informed by an indirect comparison **(Online supplement 6)**. Based on these results, CPAP may decrease IMV (RR 0.69, 95% CI 0.43 – 1.09), hospital (1.67 days fewer, 95% CI 5.43 fewer – 2.09 more) and ICU LOS (1.02 days fewer, 95% CI 3.97 days fewer – 1.93 more)(low quality evidence) but results for mortality are very uncertain (RR 0.95, 95% CI 0.52 – 1.71)). The RCT did not include children or pregnant women.

## Discussion

This rapid systematic review followed a transparent and robust approach to locate and appraise evidence from five RCTs comparing non-invasive ventilation strategies in hospitalized patients with severe or critical COVID-19 and acute hypoxemic respiratory failure not needing emergent intubation. Additional evidence was considered from 22 RCTs of similar patients without COVID-19 who had severe respiratory distress syndrome and acute respiratory failure. Based on the available evidence for mortality, use of IMV, and hospital or ICU LOS, it may be appropriate to consider the use of HFNO, NIV or CPAP instead of standard oxygen therapy for patients with severe and critical COVID-19 who do not require emergent intubation. None of the evidence for the outcomes of interest was assessed to be high quality using GRADE and ranged from moderate to very low. Additional high-quality trials are required to reduce the uncertainty associated with the evidence base for the available non-invasive ventilation strategies and to eliminate knowledge gaps related to the overall generalizability of the evidence.

This data supplements two existing evidence syntheses focused only on HFNO^28,29^ and one living systematic review^25-29^ on ventilation techniques^25-27,64^. Neither a Cochrane systematic review by Lewis et al. (2021)^28^ focused on HFNO in adult ICU patients, or a rapid systematic review by Agarwal et al. (2020) include any relevant RCTs of patients with COVID-19. None of the previous reviews include the RECOVERY-RS and HiFLO-COVID RCTs, which have been recently published. The results in this review support conclusions by Ute Muti-Schünemann et al. (2022) that HFNO may reduce mortality compared with standard oxygen therapy, and the additional RCTs included contribute to reduced uncertainty around these effects^64^. Conclusions are similar across reviews for helmet NIV compared to CPAP, owing to the availability of a single RCT in this patient population. In addition to the ongoing studies identified in this rapid review, Ute Muti-Schünemann et al. highlight other expected trials on NIV that should be watched for results in the near future: COVID-NIV *[Noninvasive Ventilation in Moderate-to-severe COVID-19-associated Acute Respiratory Distress-syndrome; NCT04667923]*, Helmet-COVID *[Helmet Non-Invasive Ventilation for COVID-19 Patients; NCT04477668]*^*65*^, PAP-COVID *[Early CPAP in COVID-19 Confirmed or Suspected Patients; NCT04390191]*, COVID HELMET *[Helmet CPAP Versus HFNC in COVID-19, NCT04395807]*, and COVID-HIGH *[HFNO vs. SOT in COVID-19; NCT04655638*]^64^. Of the ongoing studies, COVID-NIV is a case-only observational study, and PAP-COVID and COVID HELMET are listed as terminated, one due to declining cases of COVID-19, and the other citing feasibility. Two RCTs (COVID-HIGH and HELMET-COVID) are still expected to produce results in 2022 or 2023. The COVID-HIGH study (NCT NCT04655638), which has completed enrollment and will provide additional outcome data for HFNO compared to standard oxygen therapy and potentially evidence to support the role of HFNO reducing the need for escalation to NIV or CPAP (n=364 participants). The HELMET-COVID trial will contribute evidence for helmet NIV compared to standard oxygen therapy, facemask NIV, HFNO. This will provide much-needed data on the use of NIV in patients aged 14 or older with COVID-19 in the ICU and potentially comparative data for the role of NIV mask or helmet interfaces. Enrollment was reported to have been completed in October of 2021.

There are several limitations in the included RCTs that should be considered alongside the findings. There was no evidence located for use of non-invasive ventilation strategies in infants or children or important subpopulations (e.g., pregnant women), very limited evidence to support the specific use of named interfaces for the ventilation strategies considered (e.g., nasal cannula, simple face mask, venturi mask, oronasal, helmet or other), and only one RCT reported patient-important outcomes. The hospital LOS data are challenging to interpret as competing risk for death may not have been appropriately accounted for in most RCTs, or it is not possible to assess how competing risk was handled in an RCT^66^. In the set of included RCTs, the LOS outcomes are generally either secondary or exploratory outcomes, and as such, the estimates presented may be confounded by death. The LOS data for survivors and non-survivors is rarely presented. Patient heterogeneity was noted by Perkins et al. in the pragmatic RECOVERY-RS RCT but not explored in-depth^41^.

Owing to the rapid synthesis approach, there was no formal extraction or synthesis of harm outcomes. When non-invasive ventilation supports are considered, potential benefits must be consiered alongside any conceivable harms. Use of ventilation strategies could delay other clinical intervention that could lead to worsening patient prognosis, cause discomfort to the patient during use, or result in surficial wounds or pressure sores where there is skin contact.

Care for acutely ill patients with COVID-19 is complex. Although focused on ventilation strategies, the trials included likely use these interventions in conjunction with a number of other care strategies and supports (e.g., patient in prone position, medications) that should also be noted, but were not considered in detail in this rapid review. Data were insufficient to assess the impact of intervention crossover in the included RCTs which has been flagged as a concern in previous syntheses^64^. No studies specified patient outcomes or source of infection associated with any particular SARS-COV-2 variants of concern. It has been suggested that use of non-invasive ventilation strategies may increase transmission of SARS-COV-2 to healthcare workers, placing them at risk during care and adding strain to the healthcare system when isolation is required^2,26^. Although appropriate personal protective equipment may provide adequate protection for some healthcare workers when NIV strategies are used, existing studies are predominantly from early 2020 there is a lack of comparative studies available on this topic^67-78^.

The implementation of non-invasive ventilation strategies in the hospital environment must be supported by appropriate resources (including staff training, monitoring capacity) and infrastructure (e.g., interfaces or tubing, oxygen supply, cleaning or protective practices). This could prove to be challenging in some settings, particularly in low and middle income countries or settings that are acutely impacted by pandemic-related surges or supply shortages. Implementation of monitoring practices seen in the existing clinical trials may not be completely generalizable or feasible to clinical settings outside of the ICU where provider-patient ratios are higher^2^.

The evidence base is expanding and additional trials and observational studies are expected to be completed later in 2022 or early 2023. It is anticipated that studies underway will provide new information for NIV use and interfaces and provide evidence for the gaps identified in the current evidence base.

Additional information related to patient important outcomes would add substantially to our knowledge and understanding of these non-invasive ventilation strategies.

## Conclusions

This rapid systematic review highlights the available evidence to support the use of non-invasive ventilation strategies including high flow nasal oxygen, non-invasive ventilation (e.g., BiPAP) or CPAP in hospitalized patients with severe or critical COVID-19 and acute hypoxemic respiratory failure who do not need emergent intubation. Findings based on moderate to very low certainty evidence suggest that non-invasive ventilation may be considered as an alternative to standard oxygen therapy to reduce hypoxemia and dyspnea. Additional high quality RCTs are warranted to reduce uncertainty and to fill in important knowledge gaps.

## Supporting information

2022 Kelly and Wells NIV SUPPLEMENT

2022 Kelly and Wells NIV PRISMA

## Data Availability

All data produced in the present study are available upon reasonable request to the authors.

## Acknowledgements

The authors would like to acknowledge Vijay Shukla, Shuching Hsieh, and Nathan Orr for their contributions to this rapid review. We would like to acknowledge Becky Skidmore for her assistance with the search strategy design and execution. We would like to acknowledge Michael Wilson for his assistance with records from the COVID-End database.

We would also like to acknowledge our patient/citizen partners Elaine Zibrowski and Maya Sterne for their review and feedback on our protocol and their ongoing support for knowledge translation activities.

## Abbreviations

AHRF: acute hypoxemic respiratory failure
ARDS: acute respiratory distress syndrome
BiPAP: bilevel positive airway pressure
CPAP: continuous positive airway pressure
HFNC: high flow nasal cannula
HFNO: high flow nasal oxygen
IMV: invasive mechanical ventilation
MA: meta-analysis
NIV: non-invasive mechanical ventilation
NMA: network meta-analysis
NPPV: negative positive pressure ventilation
ROB: risk of bias
RCT: randomized controlled trial
RR: rapid review
SOT: standard oxygen therapy
SR: systematic review
WHO: World Health Organization

