## Supplementary material for "Noninvasive ventilation strategies for patients with severe or critical COVID-19: A rapid review of clinical outcomes": 2022 Kelly and Wells NIV SUPPLEMENT

### ONLINE SUPPLEMENTAL CONTENT

**Online supplement 1: Secondary population methods and results**

Table S1 Indirect PICO

Table S2 Methods summary

Tables S3/S4 Indirect PICO study and participant characteristics

Tables S5 to S10 indirect PICO SoF

Figure S1 indirect PICO PRISMA flow diagram

**Online supplement 2: Search strategies**

**Online supplement 3: Summary of included SR/RRs and Table**

Table S11. Systematic and rapid reviews used to identify relevant RCTs

**Online supplement 4: Detailed study and participant characteristics COVID RCTs**

Table S12 Study and participant characteristics included COVID19 RCTs

**Online supplement 5: Detailed RoB COVID RCTs**

Figure S2. RoB Summary

Tables S13 to S17. Risk of bias summary for included COVID-19 RCTs

**Online supplement 6: Detailed outcome tables**

Tables S18 to S29. Outcome tables

Table S30. Indirect data calculations

Table S31. Meta-median results for LOS outcomes - pooled

Table S32. Meta-median results for LOS outcomes - absolute

#### Online Supplement 1: Secondary PICO – ARDS and AHRF - Methods and Results

##### Research Question

In patients with acute respiratory distress syndrome (ARDS) and acute hypoxemic respiratory failure (AHRF), to what extent does high flow nasal oxygen (HFNO), continuous positive airway pressure (CPAP) or non-invasive ventilation (NIV) impact the need for invasive mechanical ventilation (IMV), hospital length of stay and death compared to standard oxygen therapy (SOT) or against each other?

##### Methods overview

Due to the uncertainty in the randomized controlled trial (RCT) evidence in severe or critical COVID-19 populations, we completed an additional rapid evidence review for non-invasive ventilation strategies in non-COVID patients with ARDS and AHRF. We implemented the population, intervention, comparator, outcomes (PICO) framework to formulate the research question (Table S1).

*Table S1: PICO framework*

| | |
|---|---|
| **Population** | Patients hospitalized with acute respiratory distress syndrome and acute hypoxemic respiratory failure that do not require emergent intubation[a] |
| **Intervention** | • High flow nasal oxygen<br>• Continuous positive airway pressure<br>• Noninvasive ventilation via facemask (or other non-helmet interfaces including nasal, oronasal, and full facial mask)<br>• Noninvasive ventilation via helmet |
| **Comparators** | Standard of care (conventional oxygen therapy) or any other intervention |
| **Outcomes** | **Primary:** Mortality (within 30, 60, 90 days, and longer if data available), need for invasive mechanical ventilation, hospital length of stay<br>**Secondary:** ICU length of stay<br>**Patient-identified outcomes of interest:** Patient comfort, satisfaction with care |
| **Eligible study designs** | Systematic/rapid reviews [b] to identify eligible trials, randomized controlled trials[c] |

a-patients weaned off IMV or who require respiratory support following IMV are not in scope.
b-eligible SR/RRs had to directly address ventilation support for two or more interventions/comparators in the PICO.
c-eligible RCTs had to directly compare two or more interventions/comparators in the PICO and at least one outcome.

We followed a similar rapid evidence review approach as for hospitalized patients with severe or critical COVID-19 and AHRF, with differences summarized below in Table S2.

*Table S2: Methods summary – Differences from direct PICO*

| | |
|---|---|
| **Search (Systematic reviews/rapid reviews)**<br><br>***May 18, 2021*** | Systematic reviews/rapid reviews used to identify relevant randomized controlled trials<br><br>A targeted search of meta-databases<br>• Epistemonikos database[1] of systematic reviews for health decision-making (includes Cochrane reviews)<br>• Living Overviews of Evidence (L.OVE) Platform |
| **Search (randomized controlled trials)**<br><br>*May 19, 2021* | Top-up of recent randomized controlled trials published since date of last systematic/rapid review search<br>• Clinicaltrials.gov<br>• International Clinical Trials Registry Platform[a]<br>• Cochrane CENTRAL<br><br>Monthly alerts continued through Dec 29, 2021.<br><br>*Date of latest included systematic review search in included randomized controlled trials used for top-up: December 1, 2020* |

a: Planned but not executed due to availability of the database. COCHRANE CENTRAL searched instead as a post hoc study registry substitution.

[1] https://www.epistemonikos.org/en/about_us/methods

#### Results

We located 22 completed randomized controlled trials (RCTs)[1-22] in 24 reports[1-24] of non-invasive ventilation support in hospitalized patients with acute respiratory distress syndrome (ARDS) and acute hypoxemic respiratory failure (AHRF) not requiring emergent intubation.

This evidence was collected using the included study lists of four systematic reviews (SRs)[25-29]. A top-up search of study registry databases found no additional eligible RCTs.

Complete results for clinical outcomes are presented in the rapid evidence report and the available evidence for non-invasive ventilation strategies for the indirect PICO is summarized using Summary of Findings tables.

##### *Identified systematic reviews*

We identified four relevant SRs (included in 7 published reports)[25-28,30-32].

1. ***Ferreyro et al. 2020***[27,31,32] completed a systematic review and network meta-analysis (NMA) examining noninvasive oxygenation strategies in adults with AHRF with a focus on mortality and intubation outcomes. "Studies that were primarily focused on the treatment of acute exacerbations of chronic obstructive pulmonary disease (i.e., >50% of the study population) or congestive heart failure (i.e., >50% of the study population) and those evaluating noninvasive oxygen strategies in the immediate post-extubation period and after major cardiovascular surgery were excluded"[27]. Methods were based on accepted SR approaches that were published in a protocol prior to execution. Limitations in the SR approach, as identified using the AMSTAR2 tool, include an unclear rationale for certain aspects of the methodology, not reporting an excluded study list and the assessment of publication bias. Methodology related to the NMA was not assessed. The search in this review is current to April 2020. A total of 25 RCTs were included. Most included RCTs compared facemask NIV to SOT (n=14), and not all included studies reported both mortality and intubation outcomes. Other included RCTs compared helmet NIV or HFNO to SOT or to each other; however, the RCTs comparing active interventions was limited. In this review, CPAP was pooled with noninvasive ventilation for all outcome comparisons. Results based on indirect comparisons showed a reduction in risk of death of endotracheal intubation with NIV strategies compared to SOT. Authors highlight the potential benefits of delivering NIV through a helmet interface, although low certainty should be considered when interpreting the results as findings are based on limited evidence. No differences in the hospital or ICU LOS were noted for any intervention.

2. ***Yasuda et al. 2021***[25] conducted a SR and NMA of noninvasive respiratory support in acute respiratory failure with a focus on associations between short-term mortality and intubation rates. A protocol was registered in advance (CRD42020139105). The review included studies of noninvasive positive pressure ventilation (NPPV), HFNO and SOT, with BiPAP and CPAP included in the NIV intervention group for syntheses. Standard SR and NMA methodology were used, with limitations noted in the December 2020 search (no alternatives to database and study registry searching), unclear data extraction methodology, inclusion of only English or Japanese language studies and no reporting of publication bias assessment. Limited study characteristics were reported, and no excluded study references were provided with reasons. The funding of studies was not investigated. Methodology related to the NMA was not assessed. A total of 25 RCTs were included. The final analysis included 19 RCTs comparing NPPV to SOT, seven comparing HFNC and SOT and five comparing HFNC and NPPV. Differences in the number of included studies and partial overlap with the Ferreyro SR/NMA are due to the fact that this

SR included studies of patients with CHF and >50% COPD while excluding studies of cardiac or abdominal surgery. This contributed to differences in findings for major outcomes compared to Ferreyro et al. and increased heterogeneity significantly in the NMA.

3. ***Baldomero et al. 2021***[28,30] conducted a SR on the effectiveness and harms of HFNO for acute respiratory failure. Standard SR methodology was used, and a protocol was registered in advance (CRD42019146691). Methods were briefly presented, but multiple bibliographic databases were searched up to July 2020. Interventions of interest were HFNO, SOT, NIV, and both pre- and post-extubation studies were included. Limitations of this SR were that the authors only included English-language studies and that methods were insufficient to conduct a fulsome assessment. A total of 29 RCTs (in 32 records) were included. Results indicated that HFNO may make little or no difference in all-cause mortality, intubation or hospital LOS compared to SOT, and data for ICU LOS is uncertain (in populations using interventions for initial management). Compared with NIV, HFNO may reduce intubation, all-cause mortality and improve patient comfort in initial acute respiratory failure management.

4. ***Lewis et al. 2021***[26] conducted a Cochrane Systematic Review using best practice methods for SRs[26]. The review updated a previously published Cochrane review that compared the use of HFNO to other types of NIV (SOT, NIV, or NIPPV, or BiPAP and CPAP) in adults requiring support to breathe in an ICU. Patients were eligible for inclusion if implemented in the ICU setting, and the patients included required respiratory support. Both pre and post-extubation RCTs were included in this review. A total of 31 RCTs were included that evaluated HFNC, NIV or CPAP. This review concluded that "HFNC may lead to less treatment failure when compared to standard oxygen therapy, but probably makes little or no difference to treatment failure when compared to NIV or NIPPV. For most other review outcomes, we found no evidence of a difference in effect. However, the evidence was often of low or very low certainty."

No additional SRs were located using monthly search alerts (current to December 29, 2021).

##### *Assessment of randomized controlled trial eligibility*

After screening all individual RCTs included in the SRs (n=74), a total of 22 RCTs (in 24 reports)[1-24] matching our indirect PICO were included. Results from the syntheses and GRADE assessments from individual SRs could not be used for mortality, IMV, and hospital or ICU LOS as a number of studies were not relevant to this PICO. Results for individual RCTs of interest were not well-reported in the SRs, and so outcome data from each study was extracted de novo. Participant and study characteristics and ROB were carried forward where possible and supplemented through the extraction of additional relevant information.

RCTs identified from the SRs were excluded if they:

a) were post-extubation or weaning interventions;
b) contained ≥ 50% participants with COPD, abdominal or cardiac surgery, or CHF;
c) did not report an outcome of interest.

##### *Results from the top-up search*

A top-up search for literature published between 1 Dec 2020 and 1 June 2021, identified a total of 1926 records. No additional RCTs were eligible for inclusion. No additional RCTs were located using monthly search alerts (current to December 29, 2021).

##### *Evidence from identified randomized controlled trials*

Twenty-two RCTs reported in 24 records were identified[1-24]. Details on study and participant characteristics and outcome data reported were extracted from the 22 RCTs identified (Tables S3 and S4). Where appropriate and feasible, data were synthesized. Where few RCTs reported mortality outcomes of interest, the longest reported mortality data were synthesized as exploratory post hoc outcomes. Results were used to inform the Summary of Findings tables (Tables S5 to S10).

The risk of bias for each trial for mortality and IMV were carried forward from the SR. None of the SRs assessed risk of bias associated with LOS outcomes as these outcomes were generally secondary or exploratory outcomes. Mortality and intubation/invasive mechanical intubation outcomes were considered in the assessment of blinding at the participant and personnel level, and intubation specifically was the specific consideration when blinding of outcome assessment was considered. SRs differed in the way they rated risk of bias due to lack of blinding (unclear or high).

Hospital LOS data are difficult to interpret as competing risk for death may not have been appropriately accounted for in most RCTs. LOS outcomes are generally secondary or exploratory outcomes in the RCTs, and as such, all indications are that estimates are confounded by death, and LOS data for survivors and non-survivors are rarely presented separately. Data were insufficient to synthesize results for hospital or ICU LOS by survivors or non-survivors in this rapid evidence review.

##### *Patient-important outcomes from SRs*

Ferreyro et al.[27] planned to synthesize meaningful results for prespecified secondary outcomes of patient comfort, but outcomes were only available in 28% of included studies and no syntheses or descriptive results were presented.

In Baldomero et al.[28] patient comfort outcomes based on percentage improved or VAS were reported in two included RCTs (872 participants), however patient populations were not relevant to the PICO as participants had COPD or were post-cardiothoracic surgery. Results in the reported evidence tables suggested that HFNO may make little or no difference in patient comfort.

Lewis et al.[26] found no evidence of a difference in comfort according to the type of respiratory support used, although this conclusion is based on some RCTs not relevant to the PICO for this rapid evidence report.

Yasuda et al.[25] did not include any patient-reported outcomes.

*Figure S1. Modified PRISMA 2020 flow diagram for identification of RCTs of ARDS and AHRF population*

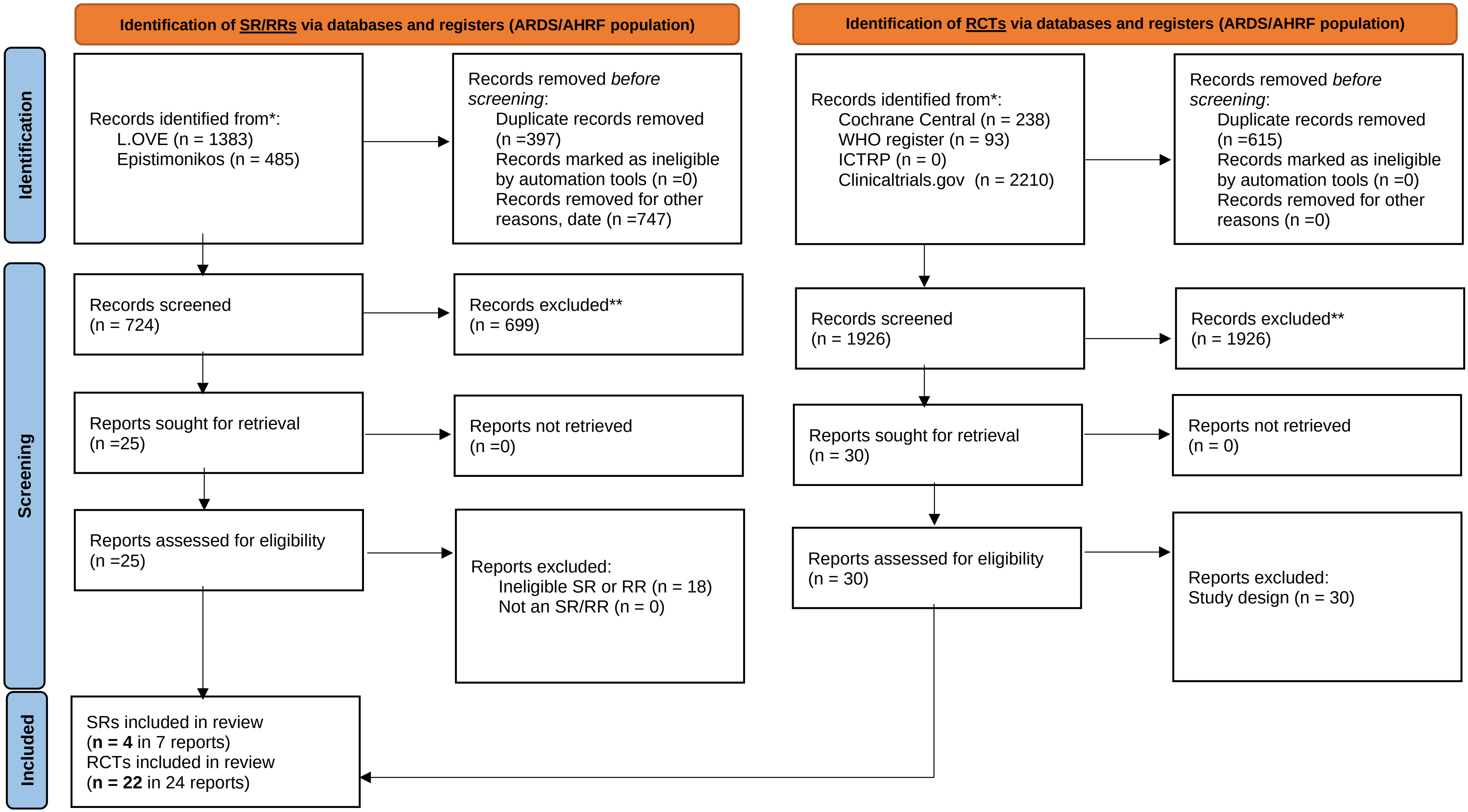


*From:* Page MJ, McKenzie JE, Bossuyt PM, Boutron I, Hoffmann TC, Mulrow CD, et al. The PRISMA 2020 statement: an updated guideline for reporting systematic reviews. BMJ 2021;372:n71. doi: 10.1136/bmj.n71. For more information, visit: http://www.prisma-statement.org/

*Table S3: Indirect PICO RCT study characteristics*

| Study | N | Intervention | Comparator 1 | Comparator 2 | Outcomes reported | | | | Study funding | Overall ROB for all-cause mortality* | Overall ROB for IMV* |
|---|---|---|---|---|---|---|---|---|---|---|---|
| | | | | | Mortality | IMV | Hospital LOS | ICU LOS | | | |
| Andino et al. 2020[1] | 46 | HFNC (n=24) | Standard oxygen (n=22) | NA | Y | Y | Y | Y | Spanish Ministry of Health, Social Services, and Equality | Low | High |
| Antonelli et al, 2000[2] | 40 | Face mask noninvasive ventilation (n = 20) | Standard oxygen (n = 20) | NA | Y | Y | Y | Y | Undisclosed | Low | Unclear |
| Azevedo et al., 2015[3] | 67 | High-flow nasal oxygen (n = 14) | Face mask noninvasive ventilation (n = 16) | NA | N | Y | N | N | Undisclosed | Unclear, **abstract only | Unclear, **abstract only |
| Azoulay et al, 2018[4] | 776 | High-flow nasal oxygen (n = 388) | Standard oxygen (n = 388) | NA | Y | Y | Y | Y | French Ministry of Health | Low | Unclear |
| Brambilla et al, 2014[5] | 81 | Helmet CPAP (n=40) | Standard oxygen (n = 41) | NA | Y | Y | Y | N | IRCCS Fondazione Ca'Granda, Ospedale Maggiore Policlinico, Milan | Unclear | High |
| Confalonieri et al, 1999[6] | 56 | Face mask noninvasive ventilation (n = 28) | Standard oxygen (n = 28) | NA | Y | Y | Y | Y | Undisclosed | Low | Unclear |
| Cosentini et al, 2010b[7] | 47 | Helmet CPAP(n = 20) | Standard oxygen (n = 27) | NA | Y | Y | N | N | Undisclosed | Low | Unclear |
| Delclaux et al, 2000[8] | 123 | Face mask CPAP (n = 62) | Standard oxygen (n = 61) | NA | Y | Y | Y | Y | Vital Signs Inc | Low | Unclear |

| Study | N | Intervention | Comparator 1 | Comparator 2 | Outcomes reported | | | | Study funding | Overall ROB for all-cause mortality* | Overall ROB for IMV* |
|---|---|---|---|---|---|---|---|---|---|---|---|
| | | | | | Mortality | IMV | Hospital LOS | ICU LOS | | | |
| Ferrer et al, 2003[9] | 105 | Face mask noninvasive ventilation (n = 51) | Standard oxygen (n = 54) | NA | Y | Y | Y | Y | Red GIRA, Red Respira, and Carburos Metalicos SA | Low | Unclear |
| Frat et al, 2015[10,23] | 310 | HFNO (n=106) | Face mask noninvasive ventilation (n = 110) | Standard oxygen (n = 94) | Y | Y | N | Y | French Ministry of Health | Low | Unclear |
| Hernandez et al, 2010[12] | 50 | Face mask noninvasive ventilation (n = 25) | Standard oxygen (n = 25) | NA | Y | Y | Y | Y | Consejería de Sanidad de Castilla | Low | Unclear |
| He et al, 2019[11] | 200 | Face mask noninvasive ventilation (n = 102) | Standard oxygen (n = 98) | NA | Y | Y | Y | Y | National Natural Science Foundation of China | High | High |
| Hilbert et al, 2001[13] | 52 | Face mask noninvasive ventilation (n = 26) | Standard oxygen (n = 26) | NA | Y | Y | N | Y | Undisclosed | Low | Unclear |
| Jones et al, 2016[14] | 303 | HFNO (n=165) | Standard oxygen (n = 138) | NA | Y | Y | Y | N | Greenlane Research and Education Fund | High | High |
| Lemiale et al, 2015[16] | 374 | Face mask noninvasive ventilation (n = 191) | Standard oxygen (n = 183) | NA | Y | Y | Y | Y | Legs Poix (Chancellerie des Universités de Paris) and OUTCOMEREA Study Group | Low | Unclear |
| Lemiale et al, 2015[2h][15] | 100 | HFNO (n=52) | Standard oxygen (n = 48) | NA | N | Y | N | N | Fisher & Paykel | High | High |

| Study | N | Intervention | Comparator 1 | Comparator 2 | Outcomes reported | | | | Study funding | Overall ROB for all-cause mortality* | Overall ROB for IMV* |
|---|---|---|---|---|---|---|---|---|---|---|---|
| | | | | | Mortality | IMV | Hospital LOS | ICU LOS | | | |
| Patel et al, 2016[17,24] | 83 | Helmet NIV (n=44) | Face mask NIV (n = 39) | NA | Y | Y | Y | Y | National Institutes of Health/National Heart, Lung, and Blood Institute | Low | Unclear |
| Shebl et al. 2018[18] | 70 | NPPV (n=36) | HFNC (n=34) | NA | Y | Y | N | N | Nil. | Unclear | Unclear/Probably High |
| Squadrone 2010[19] | 40 | Helmet CPAP (n = 20) | Standard oxygen (n = 20) | NA | Y | Y | N | N | Regione Piemonte (CEP AN RAN 07) and Ministero dell'Università (PRIN RANI 07) | Low | Unclear |
| Wermke et al., 2012[20] | 86 | Face mask noninvasive ventilation (n = 42) | Standard oxygen (n = 44) | NA | Y | Y | N | N | Undisclosed | Unclear | High |
| Wysocki et al., 1995[21] | 41 | Face mask noninvasive ventilation (n = 21) | Standard oxygen (n = 20) | NA | Y | Y | N | Y | Undisclosed | Low | Unclear |
| Zhan et al., 2012[22] | 40 | Face mask noninvasive ventilation (n = 21) | Standard oxygen (n = 19) | NA | Y | Y | Y | Y | Beijing Municipal Science and Technology Commission Program | Low | Unclear |

*Risk of bias assessment by outcome extracted from original systematic review

*Table S4: Indirect PICO RCT participant characteristics*

| Study | N | Main baseline risk factor | Main exposure | Comparator 1 | Comparator 2 | Age, mean, y | $PaO_2/FiO_2$ ratio | Respiratory rate, /min |
|---|---|---|---|---|---|---|---|---|
| Andino et al. l. 2020[1] | 46 | AHRF (pneumonia [62%]) | HFNC (n=24) | Standard oxygen (n=22) | NA | HFNC: 58 (19) COT: 61 (11) | HFNC: 96 (29) COT: 95 (37) | NR |
| Antonelli et al, 2000[2] | 40 | Mixed ARF [immunocompromised (100%)] | Face mask noninvasive ventilation (n = 20) | Standard oxygen (n = 20) | NA | 45 | 129 | 38 |
| Azevedo et al, 2015[3] | 67 | CAP (CHF [43%]) | High-flow nasal oxygen (n = 14) | Face mask noninvasive ventilation (n = 16) | NA | median 64 | NR | NR |
| Azoulay et al, 2018[4] | 776 | CAP [immunocompromised (100%)] | High-flow nasal oxygen (n = 388) | Standard oxygen (n = 388) | NA | median 64 | 132 | 33 |
| Brambilla et al, 2014[5] | 81 | CAP [immunocompromised (32%)] | Helmet CPAP (n=40) | Standard oxygen (n = 41) | NA | 67 | 141 | 34 |
| Confalonieri et al, 1999[6] | 56 | CAP | Face mask noninvasive ventilation (n = 28) | Standard oxygen (n = 28) | NA | 64 | 175 | 37 |
| Cosentini et al, 2010b[7] | 47 | CAP | Helmet CPAP (n = 20) | Standard oxygen (n = 27) | NA | 69 | 248 | 27 |
| Delclaux et al., 2000[8] | 123 | CAP | Face mask CPAP (n = 62) | Standard oxygen (n = 61) | NA | Median 58 | 144 | 33 |
| Ferrer et al., 2003[9] | 105 | CAP (immunocompromised [20%]; CHF [28%]) | Face mask noninvasive ventilation (n = 51) | Standard oxygen (n = 54) | NA | 62 | 103 | 37 |

| Study | N | Main baseline risk factor | Main exposure | Comparator 1 | Comparator 2 | Age, mean, y | $PaO_2/FiO_2$ ratio | Respiratory rate, /min |
|---|---|---|---|---|---|---|---|---|
| Frat et al, 2015[10,23] | 310 | CAP [immunocompromised (26.5%)] | HFNO (n=106) | Face mask noninvasive ventilation (n = 110) | Standard oxygen (n = 94) | 60 | 155 | 33 |
| Hernandez et al, 2010[12] | 50 | Chest trauma | Face mask noninvasive ventilation (n = 25) | Standard oxygen (n = 25) | NA | 43 | 109 | NR |
| He et al, 2019[11] | 200 | CAP | Face mask noninvasive ventilation (n = 102) | Standard oxygen (n = 98) | NA | 55 | 231 | 25 |
| Hilbert et al, 2001[13] | 52 | CAP [immunocompromised (100%)] | Face mask noninvasive ventilation (n = 26) | Standard oxygen (n = 26) | NA | 49 | 139 | 36 |
| Jones et al., 2016[14] | 303 | Mixed ARF (COPD [23.9%]; CHF [12.3%]) | HFNO (n=165) | Standard oxygen (n = 138) | NA | 73 | NR | 33 |
| Lemiale et al, 2015[16] | 374 | Pneumonia [immunocompromised (100%)] | Face mask noninvasive ventilation (n = 191) | Standard oxygen (n = 183) | NA | median 63 | 142 | 26 |
| Lemiale et al, 2015[2h][15] | 100 | Mixed ARF [immunocompromised (100%)] | HFNO (n=52) | Standard oxygen (n = 48) | NA | median 62 | 114 | 27 |
| Patel et al, 2016[17,24] | 83 | CAP [immunocompromised (100%)] | Helmet NIV (n=44) | Face mask NIV (n = 39) | NA | median 60 | 131 | 28 |
| Shebl et al. 2018[18] | 70 | AHRF (interstitial lung disease [100%]) | NPPV (n=36) | HFNC (n=34) | NA | NPPV: 61 (12) HFNC: 61 (12) | NPPV: 166 (42) HFNC: 178 (55) | NPPV: 30.1 (5.2) HFNC: 31.3 (4.8) |

| Study | N | Main baseline risk factor | Main exposure | Comparator 1 | Comparator 2 | Age, mean, y | $PaO_2/FiO_2$ ratio | Respiratory rate, /min |
|---|---|---|---|---|---|---|---|---|
| Squadrone 2010[19] | 40 | Mixed ARF (hematologic malignancies [100%]) | Helmet CPAP (n = 20) | Standard oxygen (n = 20) | NA | 49 | 269 | 30 |
| Wermke et al., 2012[20] | 86 | CAP (immunocompromised [100%]) | Face mask noninvasive ventilation (n = 42) | Standard oxygen (n = 44) | NA | median 52 | 270 | NR |
| Wysocki et al., 1995[21] | 41 | CAP (CHF [30%]) | Face mask noninvasive ventilation (n = 21) | Standard oxygen (n = 20) | NA | 63 | 207 | 35 |
| Zhan et al., 2012[22] | 40 | ALI (immunocompromised [30%]) | Face mask noninvasive ventilation (n = 21) | Standard oxygen (n = 19) | NA | 46 | 230 | 20 |

**Summary of Findings tables for the indirect PICO**

**HNFO vs SOT[1,4,10,14,15,23]**

*Table S5: Summary of Findings table for HFNO compared to SOT (indirect PICO)*

| Outcome | Study results and measurements | Absolute effect estimates SOT | Absolute effect estimates HFNO | Certainty of the Evidence (Quality of evidence) | Plain language summary |
|---|---|---|---|---|---|
| Mortality[1] | Relative risk**: 0.98 (CI 95% 0.83 - 1.15)**<br>Based on data from 1344 patients in 4 studies | **291** per 1000 | **285** per 1000<br>Difference: **6 fewer per 1000** (CI 95% 49 fewer - 44 more) | **Low** Due to very serious imprecision[2] | HFNO may have little or no difference on mortality |
| IMV | Relative risk**: 0.74 (CI 95% 0.56 - 0.99)**<br>Based on data from 668 patients in 4 studies | **207** per 1000 | **153** per 1000<br>Difference: **54 fewer per 1000** (CI 95% 91 fewer - 2 fewer) | **Moderate** Due to serious imprecision[3] | HFNO probably decreases IMV |
| Hospital LOS | Measured by: Scale: - Lower better<br>Based on data from 998 patients in 2 studies | **16.26** days Median | **14.46** days Median<br>Difference: **1.17 fewer** (CI 95% 3.16 fewer - 0.83 more) | **Moderate** Due to serious imprecision[4] | HFNO probably decreases hospital LOS |
| ICU LOS | Based on data from 996 patients in 2 studies | Studies were not pooled | | **Very low** Due to extremely serious inconsistency[7] | We are very uncertain of the impact of HFNO on ICU LOS |

1. Longest duration mortality data available, includes mix of in- hospital and end of study outcomes.
2. **Inconsistency: not serious.** The magnitude of statistical heterogeneity was moderate, with I^2: 44%.; **Imprecision: very serious.** Wide confidence intervals that include important benefit and harm;
3. **Imprecision: serious.** Number of patients does not meet the optimal information size;
4. **Imprecision: serious.** Wide confidence interval;
5. **Inconsistency: very serious.** The magnitude of statistical heterogeneity was high, with I^2: 85%, the direction of the effect is not consistent between the two included studies. One RCT suggested large benefit while one RCT suggested large harm (rated down by three).

**FACEMASK NIV vs SOT** [2,6,9-13,16,20-23]

*Table S6: Summary of Findings table for Facemask NIV compared to SOT (indirect PICO)*

| Outcome | Study results and measurements | Absolute effect estimates | | Certainty of the Evidence (Quality of evidence) | Plain language summary |
|---|---|---|---|---|---|
| | | **SOT** | **Facemask NIV** | | |
| IMV | Relative risk: **0.74** **(CI 95% 0.64 - 0.86)** Based on data from 1166 patients in 10 studies | **416** per 1000 | **308** per 1000 | **Moderate** Due to serious inconsistency[1] | Facemask NIV probably decreases IMV |
| | | Difference: **108 fewer per 1000** (CI 95% 150 fewer - 58 fewer) | | | |
| Mortality | Relative risk: **0.83** **(CI 95% 0.71 - 0.96)** Based on data from 1254 patients in 11 studies | **347** per 1000 | **288** per 1000 | **Moderate** Due to serious indirectness[2] | Facemask NIV probably decreases mortality |
| | | Difference: **59 fewer per 1000** (CI 95% 101 fewer - 14 fewer) | | | |
| Hospital LOS | Measured by: Scale: - Lower better Based on data from 829 patients in 6 studies | **20.51** days Median | **17.93** days Median | **Low** Due to serious inconsistency and serious imprecision.[3] | Facemask NIV may decrease hospital LOS |
| | | Difference: **2.02 fewer** (CI 95% 4.39 fewer - 0.35 more) | | | |
| ICU LOS | Measured by: Scale: - Lower better Based on data from 1152 patients in 10 studies | **9.43** days Median | **7.82** days Median | **Low** Due to serious inconsistency and serious imprecision[4] | Facemask NIV may decrease ICU LOS |
| | | Difference: **1.61 fewer** (CI 95% 3.21 fewer - 0.03 fewer) | | | |

1. **Inconsistency: serious.** The magnitude of statistical heterogeneity was high, with I^2: 57%.; **Indirectness: not serious.** RCT populations include immunocompromised, stem cell or solid organ transplant, mixed community-acquired pneumonia and AHRF patients;
2. Longest duration mortality data available, includes mix of in- hospital and end of study outcomes. **Indirectness: serious.** RCT populations include immunocompromised, stem cell or solid organ transplant, severe thoracic trauma, mixed community-acquired pneumonia and AHRF patients; **Imprecision: not serious.** 1.4% is considered an important reduction in mortality;

3. **Inconsistency: serious.** The magnitude of statistical heterogeneity was high, with I^2:55%; **Imprecision: serious.** Wide confidence interval that includes benefit and harm;
4. **Inconsistency: serious.** The magnitude of statistical heterogeneity was high, with I^2: 75%; **Imprecision: serious.** Wide confidence interval that includes benefit and harm.

**HELMET CPAP vs SOT[5,7,19]**

*Table S7: Summary of Findings table for Helmet CPAP compared to SOT (indirect PICO)*

| Outcome | Study results and measurements | Absolute effect estimates | | Certainty of the Evidence (Quality of evidence) | Plain language summary |
|---|---|---|---|---|---|
| | | **SOT** | **Helmet CPAP** | | |
| Mortality | Relative risk: **0.23 (CI 95% 0.10 – 0.55)**<br><br>Based on data from 168 patients in 3 studies | **250** per 1000 | **58** per 1000 | **Very low**<br>Due to serious indirectness and very serious imprecision[1] | We are very uncertain of the impact of helmet CPAP on mortality |
| | | Difference: **192 fewer per 1000** (CI 95% 225 fewer – 112 fewer) | | | |
| IMV | Relative risk: **0.45 (CI 95% 0.15 – 1.34)**<br><br>Based on data from 168 patients in 3 studies | **102** per 1000 | **46** per 1000 | **Very low**<br>Due to serious indirectness and very serious imprecision[2] | We are very uncertain of the impact of helmet CPAP on IMV |
| | | Difference: **56 fewer per 1000** (CI 95% 87 fewer – 35 more) | | | |
| Hospital LOS | Measured by:<br>Scale: - Lower better<br><br>Based on data from 81 patients in 1 study | **14** days Median | **14.5** days Median | **Low**<br>Due to very serious imprecision[3] | Helmet CPAP may have little or no difference on hospital LOS |
| | | Difference: **0.5 more** (CI 95% 3.75 fewer - 4.75 more) | | | |
| ICU LOS | No studies were found that looked at ICU LOS | | | | |

1. **Risk of Bias: not serious.** One trial stopped earlier than scheduled, potential for overestimating benefits; **Indirectness: serious.** One of three RCTs was in patients with hematologic malignancies; **Imprecision: very serious.** Number of patients is far less than would be required to meet the optimal information size (<25%);
2. **Inconsistency: not serious.** The magnitude of statistical heterogeneity was high, with I^2: 64%; **Indirectness: serious.** One of three RCTs was in patients with hematologic malignancies; **Imprecision: very serious.** Wide confidence interval that includes important benefit and harm;
3. **Imprecision: very serious.** Wide confidence interval that includes important benefit and harm, data from one study.

**FACEMASK CPAP vs SOT**[8]

*Table S8: Summary of Findings table for Facemask CPAP compared to SOT (indirect PICO)*

| Outcome | Study results and measurements | Absolute effect estimates | | Certainty of the Evidence (Quality of evidence) | Plain language summary |
|---|---|---|---|---|---|
| | | SOT | Facemask CPAP | | |
| Mortality | Relative risk**: 0.71 (CI 95% 0.38 - 1.32)**<br><br>Based on data from 123 patients in 1 study | **295** per 1000 | **209** per 1000 | **Very low** Due to extremely serious imprecision[1] | We are very uncertain of the impact of facemask CPAP on mortality |
| | | Difference: **86 fewer per 1000** (CI 95% 183 fewer - 94 more) | | | |
| IMV | Relative risk**: 0.86 (CI 95% 0.54 - 1.37)**<br><br>Based on data from 123 patients in 1 study | **393** per 1000 | **338** per 1000 | **Very low** Due to extremely serious imprecision[1] | We are very uncertain of the impact of facemask CPAP on IMV |
| | | Difference: **55 fewer per 1000** (CI 95% 181 fewer - 145 more) | | | |
| Hospital LOS | Measured by: Scale: - Lower better<br><br>Based on data from 81 patients in 1 study | **16** days Median | **14** days Median | **Very low** Due to extremely serious imprecision[1] | We are very uncertain of the impact of facemask CPAP on hospital LOS |
| | | Difference: **2 fewer** (CI 95% 17.5 fewer - 13.5 more) | | | |
| ICU LOS | Measured by: Scale: - Lower better<br><br>Based on data from 81 patients in 1 study | **9** days Median | **9** days Median | **Very low** Due to extremely serious imprecision[1] | We are very uncertain of the impact of facemask CPAP on ICU LOS |
| | | Difference: **0 fewer** (CI 95% 8.89 fewer - 8.89 more) | | | |

1. **Imprecision: extremely serious.** Wide confidence intervals that include important large benefit and harm (rated down by three levels); Data from one study.

**FACEMASK NIV vs HNFO[3,10,18]**

*Table S9: Summary of Findings table for Facemask NIV compared to HFNO (indirect PICO)*

| **Outcome** Timeframe | **Study results and measurements** | **Absolute effect estimates** **HFNO** | **Facemask NIV** | **Certainty of the Evidence** (Quality of evidence) | **Plain language summary** |
|---|---|---|---|---|---|
| Mortality | Relative risk: **1.83** **(CI 95% 1.15 – 2.89)** Based on data from 286 patients in 2 studies | **157** per 1000 | **287** per 1000 | **Very low** Due to serious indirectness and very serious imprecision[1] | We are very uncertain of the impact of facemask NIV on mortality |
| | | Difference: **130 more per 1000** (CI 95% 24 more - 297 more) | | | |
| IMV | Relative risk: **1.22** **(CI 95% 0.94 - 1.59)** Based on data from 316 patients in 3 studies | **364** per 1000 | **444** per 1000 | **Very low** Due to serious indirectness, serious risk of bias and serious imprecision[2] | We are very uncertain of the impact of facemask NIV on IMV |
| | | Difference: **80 more per 1000** (CI 95% 22 fewer - 215 more) | | | |
| Hospital LOS | No studies were found that looked at hospital LOS | | | | |
| ICU LOS | Measured by: Scale: - Lower better Based on data from 216 patients in 1 study | **12.8** days Median | **13.35** days Median | **Low** Due to very serious imprecision[3] | Facemask NIV may have little or no difference on ICU LOS |
| | | Difference: **0.55 more** (CI 95% 3.16 fewer - 4.26 more) | | | |

1. **Inconsistency: not serious.** The magnitude of statistical heterogeneity was moderately high, with I^2: 51%; **Indirectness: serious.** Differences between the population of interest and those studied (One of two RCTs 100% in interstitial lung disease patients, the other 100% with community-acquired pneumonia); **Imprecision: very serious.** Number of patients is far less than would be required to meet the optimal information size (<30%);
2. **Risk of Bias: serious.** Two of three trials have unclear sequence generation and concealment of allocation during randomization process (one is a research abstract with incomplete data); **Indirectness: serious.** Differences between the population of interest and those studied (One of three RCTs 100% in interstitial lung disease patients, one reports 100% with community-acquired pneumonia, and a third reports mixed acute respiratory failure and community-acquired pneumonia); **Imprecision: serious.** Wide confidence interval contains important benefit and harm;
3. **Imprecision: very serious.** Wide confidence intervals that include benefit and harm. Data from one study.

**HELMET NIV versus FACEMASK NIV[17,24]**

*Table S10: Summary of Findings table for Helmet NIV compared to Facemask NIV (indirect PICO)*

| Outcome | Study results and measurements | Absolute effect estimates | | Certainty of the Evidence (Quality of evidence) | Plain language summary |
|---|---|---|---|---|---|
| | | Facemask NIV | Helmet NIV | | |
| Mortality | Relative risk: **0.60 (CI 95% 0.37 - 0.99)**<br><br>Based on data from 83 patients in 1 study | **564** per 1000 | **338** per 1000 | **Low**<br>Due to very serious imprecision[1] | Helmet NIV may decrease mortality |
| | | Difference: **226 fewer per 1000** (CI 95% 355 fewer - 6 fewer) | | | |
| IMV | Relative risk**: 0.30 (CI 95% 0.15 - 0.58)**<br><br>Based on data from 83 patients in 1 study | **615** per 1000 | **185** per 1000 | **Low**<br>Due to very serious imprecision[1] | Helmet NIV may decrease IMV |
| | | Difference: **430 fewer per 1000** (CI 95% 523 fewer - 258 fewer) | | | |
| Hospital LOS | Measured by:<br>Scale: - Lower better<br><br>Based on data from 83 patients in 1 study | **7.8** days Median | **4.7** days Median | **Low**<br>Due to very serious imprecision[1] | Helmet NIV may decrease hospital LOS |
| | | Difference: **5.1 fewer** (CI 95% 9.38 fewer - 0.82 fewer) | | | |
| ICU LOS | No studies were found that looked at ICU LOS | | | | |

1. **Imprecision: very serious.** Number of patients is far less than would be required to meet the optimal information size (<10%).

#### Online Supplement 2: Search strategies for the primary COVID-19 and AHRF population

##### *Search for systematic and rapid reviews*

| | |
|---|---|
| **Database** | COVID-19 Global literature on coronavirus disease |
| **URL** | https://search.bvsalud.org/global-literature-on-novel-coronavirus-2019-ncov/ |
| **Search terms** | "high flow oxygen" or "high-flow oxygen" or "highflow oxygen" or "high frequency oxygen" or "high-frequency oxygen" or "high flow cannula" or "high-flow cannula" or "highflow cannula" or "high frequency cannula" or "high-frequency cannula" or "high flow cannulae" or "high-flow cannulae" or "highflow cannulae" or "high frequency cannulae" or "high-frequency cannulae" or HFNC or HFOC or "HFN oxygen" or "HFN O2" or "nasal cannula" or "nasal cannulae"<br>OR<br>"high flow nasal" or "high-flow nasal" or "highflow nasal" or "high frequency nasal" or "high-frequency nasal"<br>OR<br>NIV or FNIV or "F-NIV" or HNIV or "H-NIV"<br>OR<br>"controlled ventilation"<br>OR<br>"continuous positive airway pressure" or "continuous positive air-way pressure" or "bilevel positive airway pressure" or "bilevel positive air-way pressure" or "bi-level positive airway pressure" or "bi-level positive air-way pressure" or "biphasic positive airway pressure" or "biphasic positive air-way pressure" or "bi-phasic positive airway pressure" or "bi-phasic positive air-way pressure"<br>OR<br>CPAP or nCPAP or BiPAP<br>OR<br>Vapotherm or Vapo-therm or Optiflow or Opti-flow or "transnasal insuDlation" or "trans-nasal insuDlation" or "Ambu Res-cue mask" or "Ambu Res-cue masks" or Easyfit or Performatrack or Performax or "transnasal mask" or "transnasal masks" or "trans-nasal mask" or "trans-nasal masks"<br>OR<br>"mechanical ventilation" or "mechanical respiration" or "artificial ventilation" or "artificial respiration" or "artificial airway" or "artificial air-way" or "artificial airways" or "artificial air-ways"<br>OR<br>"high frequency ventilation" or "high-frequency ventilation"<br>OR<br>"invasive ventilation" or IMV<br>OR<br>"airway pressure release" and ventilat*<br>OR<br>APRV<br>OR<br>"positive pressure breathing" AND inspiratory<br>OR<br>"positive pressure breathing" AND intermittent<br>OR<br>IPPB<br>OR<br>"fluoro-carbon" AND ventilat*<br>OR<br>fluorocarbon AND ventilat*<br>OR<br>"standard oxygen" or "standard O2" or "conventional oxygen" or "conventional O2" or "oxygen therapy" or "O2 therapy" or "oxygen inhalation therapy" or "O2 inhalation therapy" or "enriched air"<br>OR<br>"non-invasive" and oxygenat*<br>OR<br>noninvasive and oxygenat*<br>OR<br>"non-invasive" and ventilat*<br>OR<br>non-invasive and ventilat*<br>OR<br>Intubat*<br>OR |

| | |
|---|---|
| | "endotracheal tube" or "endotracheal tubes" or "endotracheal tubation" or "endotracheal tubations" or "endotracheal ventilation" or "endo-tracheal tube" or "endo-tracheal tubes" or "endo-tracheal tubation" or "endo-tracheal tubations" or "endo-tracheal ventilation"<br>OR<br>tracheostom* OR tracheotom*<br>(tw:("high flow oxygen" or "high-flow oxygen" or "highflow oxygen" or "high frequency oxygen" or "high-frequency oxygen" or "high flow cannula" or "high-flow cannula" or "highflow cannula" or "high frequency cannula" or "high-frequency cannula" or "high flow cannulae" or "high-flow cannulae" or "highflow cannulae" or "high frequency cannulae" or "high-frequency cannulae" or HFNC or HFOC or "HFN oxygen" or "HFN O2" or "nasal cannula" or "nasal cannulae")) OR (tw:("high flow nasal" or "high-flow nasal" or "highflow nasal" or "high frequency nasal" or "high-frequency nasal")) OR (tw:(NIV or FNIV or "F-NIV" or HNIV or "H-NIV")) OR (tw:("non-invasive" and oxygenat*)) OR (tw:(noninvasive and oxygenat*)) OR (tw:("non-invasive" and ventilat*)) OR (tw:(non-invasive and ventilat*)) OR (tw:("controlled ventilation")) OR (tw:("continuous positive airway pressure" or "continuous positive air-way pressure" or "bilevel positive airway pressure" or "bilevel positive air-way pressure" or "bi-level positive airway pressure" or "bi-level positive air-way pressure" or "biphasic positive airway pressure" or "biphasic positive air-way pressure" or "bi-phasic positive airway pressure" or "bi-phasic positive air-way pressure")) OR (tw:(CPAP or nCPAP or BiPAP)) OR (tw:(Vapotherm or Vapo-therm or Optiflow or Opti-flow or "transnasal insuDlation" or "trans-nasal insuDlation" or "Ambu Res-cue mask" or "Ambu Res-cue masks" or Easyfit or Performatrack or Performax or "transnasal mask" or "transnasal masks" or "trans-nasal mask" or "trans-nasal masks")) OR (tw:("mechanical ventilation" or "mechanical respiration" or "artificial ventilation" or "artificial respiration" or "artificial airway" or "artificial air-way" or "artificial airways" or "artificial air-ways")) OR (tw:("high frequency ventilation" or "high-frequency ventilation")) OR (tw:("invasive ventilation" or IMV)) OR (tw:("airway pressure release" and ventilat*)) OR (tw:(APRV)) OR (tw:("positive pressure breathing" AND inspiratory)) OR (tw:("positive pressure breathing" AND intermittent)) OR (tw:(IPPB)) OR (tw:("fluoro-carbon" AND ventilat*)) OR (tw:(fluorocarbon AND ventilat*)) OR (tw:("standard oxygen" or "standard O2" or "conventional oxygen" or "conventional O2" or "oxygen therapy" or "O2 therapy" or "oxygen inhalation therapy" or "O2 inhalation therapy" or "enriched air")) OR (tw:(intubat*)) OR (tw:("endotracheal tube" or "endotracheal tubes" or "endotracheal tubation" or "endotracheal tubations" or "endotracheal ventilation" or "endo-tracheal tube" or "endo-tracheal tubes" or "endo-tracheal tubation" or "endo-tracheal tubations" or "endo-tracheal ventilation")) OR (tw:(tracheostom* OR tracheotom*))<br><br>Refined by:<br>Systematic Review, Evidence Synthesis, Broad Synthesis |
| **Study types** | Systematic or rapid reviews |
| **Search date** | 3 May 2021 |

| | |
|---|---|
| **Database** | L·OVE: Living OVerview of Evidence platform |
| **URL** | https://iloveevidence.com/[2] |
| **Search terms** | Prevention or Treatment>Procedures>Respiratory Support>HFNC<br>HFNC<br>11 Broad Syntheses<br>16 Systematic Reviews<br><br>Prevention or Treatment>Procedures>Respiratory Support>Mechanical Ventilation<br>6 Broad Syntheses<br>4 Systematic Reviews<br><br>Prevention or Treatment>Procedures>Respiratory Support>Breathing Gases>Oxygen<br>2 Broad Syntheses<br><br>Prevention or Treatment>Procedures>Respiratory Support>Breathing Gases>Hydrogen/Oxygen<br>No Broad Syntheses/Systematic Reviews<br><br>Prevention or Treatment>Procedures>Respiratory Support>Tracheostomy<br>28 Broad Syntheses<br>6 Systematic Reviews |
| **Study types** | Systematic or rapid reviews |
| **Search date** | 2 May 2021 |

[2] Note: The L.OVE beta interface was not working for complex queries – no results were obtained using this approach.

| | |
|---|---|
| **Database** | COVID-END Platform |
| **URL** | https://www.mcmasterforum.org/networks/covid-end/resources-to-support-decision-makers/Inventory-of-best-evidence-syntheses/clinical-management |
| **Search terms** | Scanned Clinical Treatment category<br>Total: 9 records under "Invasive ventilation" and "Non-invasive ventilation"<br><br>This item listed as a review but it is an RCT and was assessed for eligibility - https://covid-nma.com/living_data/index.php?comparison=165 |
| **Study types** | Systematic or rapid reviews |
| **Search date** | 2 May 2021 |

#### *Search for RCTs available after the search of the latest systematic/rapid reviews*

| | |
|---|---|
| **Database** | WHO COVID-19 Global literature on coronavirus disease |
| **URL** | https://search.bvsalud.org/global-literature-on-novel-coronavirus-2019-ncov/ |
| **Search terms** | "high flow oxygen" or "high-flow oxygen" or "highflow oxygen" or "high frequency oxygen" or "high-frequency oxygen" or "high flow cannula" or "high-flow cannula" or "highflow cannula" or "high frequency cannula" or "high-frequency cannula" or "high flow cannulae" or "high-flow cannulae" or "highflow cannulae" or "high frequency cannulae" or "high-frequency cannulae" or HFNC or HFOC or "HFN oxygen" or "HFN O2" or "nasal cannula" or "nasal cannulae"<br>OR<br>"high flow nasal" or "high-flow nasal" or "highflow nasal" or "high frequency nasal" or "high-frequency nasal"<br>OR<br>NIV or FNIV or "F-NIV" or HNIV or "H-NIV"<br>OR<br>"controlled ventilation"<br>OR<br>"continuous positive airway pressure" or "continuous positive air-way pressure" or "bilevel positive airway pressure" or "bilevel positive air-way pressure" or "bi-level positive airway pressure" or "bi-level positive air-way pressure" or "biphasic positive airway pressure" or "biphasic positive air-way pressure" or "bi-phasic positive airway pressure" or "bi-phasic positive air-way pressure"<br>OR<br>CPAP or nCPAP or BiPAP<br>OR<br>Vapotherm or Vapo-therm or Optiflow or Opti-flow or "transnasal insuDlation" or "trans-nasal insuDlation" or "Ambu Res-cue mask" or "Ambu Res-cue masks" or Easyfit or Performatrack or Performax or "transnasal mask" or "transnasal masks" or "trans-nasal mask" or "trans-nasal masks"<br>OR<br>"mechanical ventilation" or "mechanical respiration" or "artificial ventilation" or "artificial respiration" or "artificial airway" or "artificial air-way" or "artificial airways" or "artificial air-ways"<br>OR<br>"high frequency ventilation" or "high-frequency ventilation"<br>OR<br>"invasive ventilation" or IMV<br>OR<br>"airway pressure release" and ventilat*<br>OR<br>APRV<br>OR<br>"positive pressure breathing" AND inspiratory<br>OR<br>"positive pressure breathing" AND intermittent<br>OR<br>IPPB<br>OR<br>"fluoro-carbon" AND ventilat*<br>OR<br>fluorocarbon AND ventilat*<br>OR<br>"standard oxygen" or "standard O2" or "conventional oxygen" or "conventional O2" or "oxygen therapy" or "O2 therapy" or "oxygen inhalation therapy" or "O2 inhalation therapy" or "enriched air"<br>OR<br>"non-invasive" and oxygenat* |

| | |
|---|---|
|  | OR<br>noninvasive and oxygenat*<br>OR<br>"non-invasive" and ventilat*<br>OR<br>non-invasive and ventilat*<br>OR<br>Intubat*<br>OR<br>"endotracheal tube" or "endotracheal tubes" or "endotracheal tubation" or "endotracheal tubations" or "endotracheal ventilation" or "endo-tracheal tube" or "endo-tracheal tubes" or "endo-tracheal tubation" or "endo-tracheal tubations" or "endo-tracheal ventilation"<br>OR<br>tracheostom* OR tracheotom*<br>(tw:("high flow oxygen" or "high-flow oxygen" or "highflow oxygen" or "high frequency oxygen" or "high-frequency oxygen" or "high flow cannula" or "high-flow cannula" or "highflow cannula" or "high frequency cannula" or "high-frequency cannula" or "high flow cannulae" or "high-flow cannulae" or "highflow cannulae" or "high frequency cannulae" or "high-frequency cannulae" or HFNC or HFOC or "HFN oxygen" or "HFN O2" or "nasal cannula" or "nasal cannulae")) OR (tw:("high flow nasal" or "high-flow nasal" or "highflow nasal" or "high frequency nasal" or "high-frequency nasal")) OR (tw:(NIV or FNIV or "F-NIV" or HNIV or "H-NIV")) OR (tw:("non-invasive" and oxygenat*)) OR (tw:(noninvasive and oxygenat*)) OR (tw:("non-invasive" and ventilat*)) OR (tw:(non-invasive and ventilat*)) OR (tw:("controlled ventilation")) OR (tw:("continuous positive airway pressure" or "continuous positive air-way pressure" or "bilevel positive airway pressure" or "bilevel positive air-way pressure" or "bi-level positive airway pressure" or "bi-level positive air-way pressure" or "biphasic positive airway pressure" or "biphasic positive air-way pressure" or "bi-phasic positive airway pressure" or "bi-phasic positive air-way pressure")) OR (tw:(CPAP or nCPAP or BiPAP)) OR (tw:(Vapotherm or Vapo-therm or Optiflow or Opti-flow or "transnasal insuDlation" or "trans-nasal insuDlation" or "Ambu Res-cue mask" or "Ambu Res-cue masks" or Easyfit or Performatrack or Performax or "transnasal mask" or "transnasal masks" or "trans-nasal mask" or "trans-nasal masks")) OR (tw:("mechanical ventilation" or "mechanical respiration" or "artificial ventilation" or "artificial respiration" or "artificial airway" or "artificial air-way" or "artificial airways" or "artificial air-ways")) OR (tw:("high frequency ventilation" or "high-frequency ventilation")) OR (tw:("invasive ventilation" or IMV)) OR (tw:("airway pressure release" and ventilat*)) OR (tw:(APRV)) OR (tw:("positive pressure breathing" AND inspiratory)) OR (tw:("positive pressure breathing" AND intermittent)) OR (tw:(IPPB)) OR (tw:("fluoro-carbon" AND ventilat*)) OR (tw:(fluorocarbon AND ventilat*)) OR (tw:("standard oxygen" or "standard O2" or "conventional oxygen" or "conventional O2" or "oxygen therapy" or "O2 therapy" or "oxygen inhalation therapy" or "O2 inhalation therapy" or "enriched air")) OR (tw:(intubat*)) OR (tw:("endotracheal tube" or "endotracheal tubes" or "endotracheal tubation" or "endotracheal tubations" or "endotracheal ventilation" or "endo-tracheal tube" or "endo-tracheal tubes" or "endo-tracheal tubation" or "endo-tracheal tubations" or "endo-tracheal ventilation")) OR (tw:(tracheostom* OR tracheotom*))<br><br>Refined by: Controlled Clinical Trial, Year 2020-2021 |
| **Study types** | Randomized studies of interventions |
| **Search date** | July 2020 to 17 June 2021 (with alerts continued to Dec 2021, ongoing studies were all checked for results or status changes to same date) |

| | |
|---|---|
| **Database** | Cochrane COVID-19 study register |
| **URL** | https://covid-19.cochrane.org/ |
| **Search terms** | Intervention – Randomised – Covid 19 – Mechanical Ventilation<br>13 studies selected for export. Note: Export may contain more than one citation per study. (18 studies exported)<br><br>Intervention – Randomised – Covid 19 – HFNO<br>2 studies selected for export. Note: Export may contain more than one citation per study. (2 studies exported)<br><br>Intervention – Randomised – Covid 19 – HFNO<br>6 studies selected for export. Note: Export may contain more than one citation per study. (8 studies exported)<br><br>Intervention – Randomised – Covid 19 – Invasive Ventilation<br>2 studies selected for export. Note: Export may contain more than one citation per study. (2 studies exported)<br><br>Intervention – Randomised – Covid 19 – HFOT |

| | |
|---|---|
| | 1 studies selected for export. Note: Export may contain more than one citation per study. (2 studies exported)<br><br>Intervention – Randomised – Covid 19 – Invastive Mechanical Ventilation<br>1 studies selected for export. Note: Export may contain more than one citation per study. (1 studies exported)<br><br>Intervention – Randomised – Covid 19 – NPPV<br>1 studies selected for export. Note: Export may contain more than one citation per study. (1 studies exported)<br><br>Intervention – Randomised – Covid 19 – O2<br>1 studies selected for export. Note: Export may contain more than one citation per study. (1 studies exported)<br><br>Intervention – Randomised – Covid 19 – O2 Therapy Support<br>1 studies selected for export. Note: Export may contain more than one citation per study. (1 studies exported) |
| **Study types** | Randomized studies of interventions |
| **Search date** | Searches limited to 2020 Jul 1 – 2021 May 15, with alerts to December 29, 2021 |

| | |
|---|---|
| **Database** | Clinicaltrials.gov study register |
| **URL** | https://clinicaltrials.gov |
| **Search terms** | 596 Studies found for: ventilation \| Interventional Studies \| COVID-19 \| First posted from 07/01/2020 to 05/15/2021<br><br>187 Studies found for: cannula OR cannulae OR "high flow" OR highflow OR HFNC OR HFOC or HFNO OR "HFN Oxygen" OR "HFN O2" OR "high frequency nasal" \| Interventional Studies \| COVID-19 \| First posted from 07/01/2020 to 05/15/2021<br><br>55 Studies found for: (("non-invasive" or noninvasive) AND (oxygen OR oxygenation OR ventilation OR respiration or respiratory)) OR (NIV OR FNIV OR "F-NIV" OR HNIV OR "H-NIV") \| Interventional Studies \| COVID-19 \| First posted from 07/01/2020 to 05/15/2021<br><br>No Studies found for: "controlled ventilation" or "positive airway pressure" or "positive air-way pressure" OR CPAP or nCPAP or BiPAP \| Interventional Studies \| COVID-19 \| First posted from 07/01/2020 to 05/15/2021<br><br>No Studies found for: Vapotherm or Vapo-therm or Optiflow or Opti-flow or "transnasal insuDlation" or "trans-nasal insuDlation" or "Ambu Res-cue mask" or "Ambu Res-cue masks" or Easyfit or Performatrack or Performax or transnasal mask or trans-nasal mask \| Interventional Studies \| COVID-19 \| First posted from 07/01/2020 to 05/15/2021<br><br>No Studies found for: "high frequency ventilation" or "high-frequency ventilation" \| Interventional Studies \| COVID-19 \| First posted from 07/01/2020 to 05/15/2021<br><br>No Studies found for: "invasive ventilation" or IMV \| Interventional Studies \| COVID-19 \| First posted from 07/01/2020 to 05/15/2021<br><br>No Studies found for: "airway pressure release" and ventilat* \| Interventional Studies \| COVID-19 \| First posted from 07/01/2020 to 05/15/2021<br><br>1 Study found for: (invasive or mechanical or artificial) AND (ventilation or respiration or respiratory) \| Interventional Studies \| COVID-19 \| First posted from 07/01/2020 to 05/15/2021<br><br>No Studies found for: (invasive or mechanical or artificial) AND (airway or "air-way") \| Interventional Studies \| COVID-19 \| First posted from 07/01/2020 to 05/15/2021<br><br>No Studies found for: "airway pressure release" OR APRV \| Interventional Studies \| COVID-19 \| First posted from 07/01/2020 to 05/15/2021<br><br>No Studies found for: "positive pressure breathing" or IPPB \| Interventional Studies \| COVID-19 \| First posted from 07/01/2020 to 05/15/2021 |

| | |
|---|---|
| | No Studies found for: (fluorocarbon or fluoro carbon) AND ventilation \| Interventional Studies \| COVID-19 \| First posted from 07/01/2020 to 05/15/2021<br><br>No Studies found for: fluorocarbon or "fluoro carbon" \| Interventional Studies \| COVID-19 \| First posted from 07/01/2020 to 05/15/2021<br><br>No Studies found for: "oxygen therapy" or "O2 therapy" or "oxygen inhalation therapy" or "O2 inhalation therapy" or "enriched air" \| Interventional Studies \| COVID-19 \| First posted from 07/01/2020 to 05/15/2021<br><br>No Studies found for: oxgenation \| Interventional Studies \| COVID-19 \| First posted from 07/01/2020 to 05/15/2021<br><br>8 Studies found for: tracheostomy OR tracheotomy \| Interventional Studies \| COVID-19 \| First posted from 07/01/2020 to 05/15/2021<br><br>No Studies found for: intubate or intubation or tubation \| Interventional Studies \| COVID-19 \| First posted from 07/01/2020 to 05/15/2021 |
| **Study types** | Randomized studies of interventions |
| **Search date** | Searches limited to 2020 Jul 1 – 2021 May 15, with alerts to December 29, 2021 |

#### Search strategies for non-COVID-19 population (ARDS and AHRF)

| **Database** | Epistemonikos |
|---|---|
| **URL** | https://www.epistemonikos.org/ |
| **Search terms** | (advanced_title_en:(ventilat* OR cannula* OR HFNC OR HFOC OR "HFN oxygen" OR "HFN O2" OR NIV OR FNIV OR "F-NIV" OR HNIV OR "H-NIV" OR "positive airway pressure" OR ""positive air-way pressure" OR CPAP OR nCPAP OR BiPAP OR "high flow oxygen" OR "highflow oxygen" OR "high frequency oxygen" OR oxygenat* OR "high flow nasal" OR "high-flow nasal" OR "highflow nasal" OR "high frequency nasal" OR "transnasal mask" OR "transnasal masks" OR "trans-nasal mask" OR "trans-nasal masks" OR IMV OR "mechanical respiration" OR "artificial respiration" OR "artificial airway" OR "artificial air-way" OR "artificial airways" OR "artificial air-ways" OR "airway pressure release" OR APRV OR "positive pressure breathing" OR "standard oxygen" OR "standard O2" OR "conventional oxygen" OR "conventional O2" OR "oxygen therapy" OR "O2 therapy" OR "oxygen inhalation therapy" OR "O2 inhalation therapy" OR "enriched air" OR intubat* OR tubation* OR tube OR tubes OR tracheostom* OR tracheotom*) OR advanced_abstract_en:(ventilat* OR cannula* OR HFNC OR HFOC OR "HFN oxygen" OR "HFN O2" OR NIV OR FNIV OR "F-NIV" OR HNIV OR "H-NIV" OR "positive airway pressure" OR ""positive air-way pressure" OR CPAP OR nCPAP OR BiPAP OR "high flow oxygen" OR "highflow oxygen" OR "high frequency oxygen" OR oxygenat* OR "high flow nasal" OR "high-flow nasal" OR "highflow nasal" OR "high frequency nasal" OR "transnasal mask" OR "transnasal masks" OR "trans-nasal mask" OR "trans-nasal masks" OR IMV OR "mechanical respiration" OR "artificial respiration" OR "artificial airway" OR "artificial air-way" OR "artificial airways" OR "artificial air-ways" OR "airway pressure release" OR APRV OR "positive pressure breathing" OR "standard oxygen" OR "standard O2" OR "conventional oxygen" OR "conventional O2" OR "oxygen therapy" OR "O2 therapy" OR "oxygen inhalation therapy" OR "O2 inhalation therapy" OR "enriched air" OR intubat* OR tubation* OR tube OR tubes OR tracheostom* OR tracheotom*)) AND (advanced_title_en:((advanced_title_en:(acute respiratory distress) OR advanced_abstract_en:(acute respiratory distress)) OR (advanced_title_en:(ards) OR advanced_abstract_en:(ards)) OR (advanced_title_en:(acute hypoxemic respiratory failure) OR advanced_abstract_en:(acute hypoxemic respiratory failure)) OR (advanced_title_en:(acute hypoxaemic respiratory failure) OR advanced_abstract_en:(acute hypoxaemic respiratory failure)) OR (advanced_title_en:(AHRF) OR advanced_abstract_en:(AHRF)) OR (advanced_title_en:(shock lung) OR advanced_abstract_en:(shock lung))) OR advanced_abstract_en:((advanced_title_en:(acute respiratory distress) OR advanced_abstract_en:(acute respiratory distress)) OR (advanced_title_en:(ards) OR advanced_abstract_en:(ards)) OR (advanced_title_en:(acute hypoxemic respiratory failure) OR advanced_abstract_en:(acute hypoxemic respiratory failure)) OR (advanced_title_en:(acute hypoxaemic respiratory failure) OR advanced_abstract_en:(acute hypoxaemic respiratory failure)) OR (advanced_title_en:(AHRF) OR advanced_abstract_en:(AHRF)) OR (advanced_title_en:(shock lung) OR advanced_abstract_en:(shock lung)))) [Filters: protocol=no, classification=systematic-review] |
| **Study types** | Systematic or rapid reviews |
| **Search date** | 18 May 2021 |

RCT top-up

| | |
|---|---|
| **Database** | EBM Reviews - Cochrane Central Register of Controlled Trials |
| **URL** | https://www.wolterskluwer.com/en/solutions/ovid/evidencebased-medicine-reviews-ebmr-904 |
| **Search terms** | 1 respiratory distress syndrome, adult/ (37)<br>2 ((respiratory or respiration or lung or ventilatory) adj2 (depress* or insufficien* or fail* or deficien* or disturb* or dysfunction* or compromis*) adj3 (acute or adult)).ti,ab,kw. (1910)<br>3 (lung adj1 shock).ti,ab,kw. (10)<br>4 ARDS.ti,ab,kw. (2155)<br>5 ARDSS.ti,ab,kw. (0)<br>6 exp Respiratory Insufficiency/ (2829)<br>7 (respiratory failure adj3 hypox?emi*).ti,ab,kw. (404)<br>8 (respiratory failure adj3 hypercapni*).ti,ab,kw. (327)<br>9 AHRF.ti,ab,kw. (90)<br>10 (acute adj2 (hypoxia or hypox?emi*)).ti,ab,kw. (670)<br>11 or/1-10 [ARDS/AHRF] (6797)<br>12 Cannula/ (113)<br>13 Oxygen/ (5200)<br>14 Oxygen Inhalation Therapy/ (1164)<br>15 11 and (13 or 14) (456)<br>16 ((high-flow or highflow or high-frequency or prolong*) adj3 cannula*).ti,ab,kw. (908)<br>17 ((high-flow or highflow or high-frequency or prolong*) adj3 nasal*).ti,ab,kw. (1332)<br>18 ((high-flow or highflow or high-frequency or prolong*) adj3 (oxygen* or O2)).ti,ab,kw. (1097)<br>19 (HFNC or HFNO or HFNP or HFOC).ti,ab,kw. (561)<br>20 (("positive pressure" or "positive end-expiratory pressure") adj3 (respirat* or ventilat*)).ti,ab,kw. (2211)<br>21 continuous positive airway pressure.ti,ab,kw. (3829)<br>22 (CPAP or nCPAP).ti,ab,kw. (5110)<br>23 (airway pressure release adj3 ventilat*).ti,ab,kw. (80)<br>24 APRV.ti,ab,kw. (69)<br>25 ((inspiratory or intermittent) adj3 positive pressure breathing).ti,ab,kw. (75)<br>26 IPPB.ti,ab,kw. (69)<br>27 ((non-invasive or noninvasive) adj3 (oxygen* or ventilat*)).ti,ab,kw. (3456)<br>28 controlled ventilation.ti,ab,kw. (849)<br>29 (bi level positive airway pressure or bilevel positive airway pressure or bi-level positive airway pressure or BiPaP or NIV).ti,ab,kw. (1635)<br>30 (FNIV or F-NIV or H-NIV or HNIV).ti,ab,kw. (20)<br>31 standard oxygen.ti,ab,kw. (206)<br>32 ((low flow or low-flow or lowflow) adj2 oxygen*).ti,ab,kw. (206)<br>33 ((mask* or helmet*) adj1 (face or oxygen)).ti,ab,kw. (1826)<br>34 (Ambu Res-cue mask* or Easyfit or Performatrack or Performax or transnasal mask* or facemask* or face-mask*).ti,ab,kw. (2042)<br>35 controlled ventilation.ti,ab,kw. (849)<br>36 exp Respiration, Artificial/ (6241)<br>37 exp Ventilators, Mechanical/ (268)<br>38 ((artificial* or mechanical*) adj3 (respirat* or ventilat*)).ti,ab,kw. (15417)<br>39 artificial airway?.ti,ab,kw. (98)<br>40 ((assist* or depend* or support*) adj3 (respirat* or ventilat*)).ti,ab,kw. (5925)<br>41 ((liquid or fluorocarbon or fluoro-carbon) adj3 ventilat*).ti,ab,kw. (42)<br>42 (high-frequency adj3 ventilat*).ti,ab,kw. (569)<br>43 (invasive* adj3 (oxygen* or ventilat*)).ti,ab,kw. (3149)<br>44 [IMV.tw,kf.] (0)<br>45 or/15-44 [VENTILATION OPTIONS] (30378)<br>46 11 and 45 [ARDS/AHRF - VENTILATION OPTIONS] (3698)<br>47 (202012* or 2021*).up. (642312)<br>48 46 and 47 [UPDATE PERIOD] (1817) |
| **Study types** | Randomized studies published after the date of the last indirect PICO SR or RR search (December 1, 2020 based on included SR) |
| **Search date** | Dec 1 2020 to 1 Jun 2021 (with alerts to December 29, 2021) |

#### Online Supplement 3: Summary of indentified SR/RRs

#### Identified systematic reviews

Three SRs reported in five records were identified[26,33-36].

1. ***Schünemann et al. (2020)*** completed a living systematic review (LSR) published as a systematic review and two additional research letters reporting updated results (current to July 2020)[33-35]. No additional updates have been published. This LSR addresses multiple research questions and streams of evidence, of which their reported PICO #1 is directly relevant to the benefits and harms of ventilation techniques for coronavirus infections, including those that causing COVID-19. The LSR had a protocol registered in advance and uses recognized SR methods and comprehensively searched 21 bibliographic databases. It was rated as a methodologically rigorous systematic review following assessment with AMSTAR2. The authors' noted in their conclusions that that direct studies in COVID-19 are limited and poorly reported based mostly on observational evidence in SARS, MERS and COVID-19. The LSR (update #1) identified one completed RCT published in April 2020 that followed patients (n=72) in the Huanggang hospital in China who were randomized to HFNC (n=37) or SOT (n=35) in patients with severe COVID-19 pneumonia and acute respiratory failure[37]. Of the eight potentially relevant in-progress RCT records identified in the Schünemann et al. LSR, one additional RCT (RECOVERY-RS) is complete as of August 4, 2021, and has results available[38]. One additional RCT[39] was identified using the reference list of the RECOVERY-RS pre-print publication.

2. ***Agarwal et al. (2020)*** completed a rapid SR updating a previous SR and meta-analysis by Rochwerg et al.(2019) comparing HFNO to SOT for two unique research questions, one of which was relevant to our PICO[36]. No protocol was registered or published. Although this SR was completed in 7 days, a search of three bibliographic databases was completed (May 2020), and standard systematic review methods were utilized. The study received a moderate rating for methodological rigour using AMSTAR2, with downgrading in the rating attributable to details that were not reported in the publication pertaining to the rationale for selection criteria, not providing reasons for excluded studies, and no investigation of publication or funding biases. **This rapid** SR did not find any RCTs that directly evaluated HFNC in patients with COVID-19 or other coronavirus infections, and studies in progress were not sought or reported.

3. ***Lewis et al. (2020)*** completed a Cochrane Systematic Review using best practice methods for SRs (rated as a rigorously conducted SR following assessment with AMSTAR2)[26]. The review updated a previously published Cochrane review that compared the use of HFNO to other types of NIV (SOT, NIV, or NIPPV, or BiPAP and CPAP) in adults requiring support to breathe in an ICU. Patients with COVID-19 were not the direct focus of the SR, but RCTs of COVID-19 patients were eligible for inclusion if implemented in the ICU setting and the patients included required respiratory support. None of the 31 included studies evaluated HFNC, NIV or CPAP in patients with COVID-19. None of the ongoing studies identified (n=51) as in-progress included patients with COVID-19.

#### Identified rapid reviews

Four additional rapid reviews using a range of accepted 'rapid review' methods were identified for inclusion[40-43]. Three RRs[41-43] were completed between March and November 2020, and one was published in May 2021[40]. No RCTs directly evaluating the use of noninvasive ventilation strategies (HFNC, NIV, or CPAP) in COVID-19 patients were identified from the RRs. Most reported results were from non-randomized studies or observational cohorts. One potentially relevant ongoing RCT comparing helmet CPAP to SOT was identified, but no results were published or posted to the study registration as of December 29, 2021 (NCT04326075).

*Table S11. Systematic and rapid reviews used to identify relevant RCTs*

| Included: | Population | Interventions studied | Outcomes reported | Search date | RCTs identified | AMSTAR2 rating |
|---|---|---|---|---|---|---|
| **Systematic reviews** | | | | | | |
| Lewis et al. 2021[26], *Cochrane Systematic Review*** | Adults (16 years or older) requiring support to breathe in an ICU | HFNC compared to other types NIV Including standard oxygen therapy, NIV, or NIPPV, or (BiPAP and CPAP) | Treatment failure, in-hospital mortality (up to 90d), ICU LOS, short- and long-term patient comfort. | 17 April 2020 | 0 RCTs in COVID-19 pts<br><br>0 ongoing RCTs in COVID-19 pts | High quality |
| Schünemann et al. 2020[33-35a], *Annals of Internal Medicine* | Patients with confirmed or probable COVID-19 infection and hypoxemic respiratory failure | PICO 1:<br><br>NIV, including Bi-PAP, CPAP, and HFNC; IMV; standard oxygen therapy; or no mechanical ventilation | death, IMV hospital LOS, ICU LOS, contextual outcomes (acceptability, feasibility, resources use, effect on equity) | Latest update 11 July 2020 | 0 RCTs in base LSR<br><br>1 RCT[37] in LSR update 1<br><br>0 RCTs in LSR Update 2<br><br>1 RCT[38] identified as in-progress with results available<br><br>1 RCT[39] identified using the | High quality |

| Included: | Population | Interventions studied | Outcomes reported | Search date | RCTs identified | AMSTAR2 rating |
|---|---|---|---|---|---|---|
| | | | | | reference list of an identified RCT | |
| Agarwal et al. 2020[36]*, *Canadian Journal of Anaesthesia* | Critically ill COVID-19 patients with acute hypoxemic respiratory failure | HFNO<br>compared to standard oxygen therapy, NIV, NIPPV (CPAP, BiPAP) | Mortality, IMV hospital LOS, ICU LOS | 14 May 2020 | 0 RCTs in COVID-19 pts<br><br>Did not report RCTs in-progress | Moderate Quality<br><br>Identified as rapid but reporting brief, so assessment of quality limited |
| **Rapid reviews** | | | | | | |
| Alberta Health Services, Alberta, Canada, 2020[43] | Acute Hypoxemic respiratory failure not due to AECOPD or CHF | Noninvasive ventilation, helmet CPAP, BiPAP | any | 6 May 2020 | 0 RCTs in COVID-19 pts | Methods not reported, unable to assess |
| Swedish Agency For Health Technology Assessment and Assessment of Social Services 2020[41] | Acute respiratory failure due to coronavirus | ‘Noninvasive ventilation’<br><br>CPAP, BiPAP, NIPPV, nasal ventilation, mask ventilation[b] | effectiveness | March 2020 | 0 RCTs in COVID-19 pts | Moderate quality |

| Included: | Population | Interventions studied | Outcomes reported | Search date | RCTs identified | AMSTAR2 rating |
|---|---|---|---|---|---|---|
| New South Wales Health, Evidence Check. Australia, 2020[42] | Patients with severe Covid-19 | CPAP, BiPAP | any | 1 and 6 April 2020 | 0 RCTs in COVID-19 pts<br><br>1 RCT in-progress[d] | Methods not reported, unable to assess |
| Radovanovic et al. 2021[40] | Patients with acute respiratory failure secondary to COVID-19 pneumonia | CPAP, NIV | In-hospital mortality | 1 Nov 2020[c] | 0 RCTs<br><br>Did not report RCTs in-progress | Moderate to low quality |

AHS=Alberta Health Services; CPAP=; BiPAP=Bilevel Positive Airway Pressure;

**Note that COVID-19 pts included in a subpopulation of adult intensive care patients (the population of interest for the review).

*Update of Rochwerg et al. 2019.

a: includes two published living updates. Multiple PICOs investigated. Data represented PICO 1 relevant to this rapid evidence review.

b: interventions identified from the provided search strategy.

c: limited specific search based on NIV and CPAP only, and in-hospital mortality.

d: EC-COVID-RCT (Helmet CPAP compared to standard oxygen, planned n=900, NCT04326075).

#### Online supplement 4: Detailed study and participant characteristics COVID RCTs

*Table S12. Participant and study characteristics for COVID-19 RCTs*

| Study/Design | Population | Interventions | Outcomes reported | Age (y), Mean±SD | PaO2.FiO2 ratio | Respiratory rate, /min | Funding |
|---|---|---|---|---|---|---|---|
| **Li et al. 2020**[37]<br><br>two-arm, parallel RCT, CHINA (single centre)<br><br>N=72 | Patients with severe coronavirus pneumonia complicated with acute respiratory failure | HFNC [n=37]<br><br>Standard oxygen therapy [n=35] | Mechanical ventilation at 12 h | HFNC<br>32±6.42<br><br>SOT<br>35±4.67 | Not reported<br><br>HFNC<br>PaO2= 63.162 ±3.912 mmHg<br><br>SOT<br>PaO2=62.886 ±3.243 mmHg | Not reported | Unclear |
| **Grieco et al. 2021**[44]<br><br>HENIVOT<br><br>NCT04502576<br><br>two-arm, parallel RCT, ITALY (4 centres)<br><br>N=109 | Patients admitted to the intensive care unit with COVID-19–induced moderate to severe hypoxemic respiratory failure | Helmet NIV [n=55]<br><br>HFNO [n=54] | Intubation, 28 d<br><br>Hospital LOS<br><br>ICU LOS | median (IQR)<br><br>Helmet NIV<br>66 (57-72)<br><br>HFNO<br>63 (55-69) | Helmet NIV<br>105 (83-125)<br><br>HFNO<br>102 (80-124) | Helmet NIV<br>28 (24-32)<br><br>HFNO<br>28 (23-32) | Funded by a research grant (2017 Merck Sharp & Dohme SRL award) by the Italian Society of Anesthesia, Analgesia, and Intensive Care Medicine |
| **Perkins et al. 2021**[38]<br><br>RECOVERY-RS<br><br>ISRCTN16912075 | Hospitalized adults with acute respiratory failure due to COVID-19 deemed suitable for tracheal intubation if | CPAP [n=380]<br><br>HFNO [n=417]<br><br>Standard oxygen therapy [n=475] | Mortality, 30 d<br><br>Intubation, 30 d<br><br>Tracheal intubation | CPAP<br>56.7 ± 12.5<br><br>HFNO<br>57.6 ± 13.0<br><br>SOT | CPAP<br>131.8 ± 67.8<br><br>HFNO<br>138.5 ±87.6<br><br>SOT | CPAP<br>26.4 ± 7.5<br><br>HFNO<br>25.4 ± 7.0<br><br>SOT | Funded and prioritized as an urgent public health COVID-19 study by the |

| Study/Design | Population | Interventions | Outcomes reported | Age (y), Mean±SD | PaO2.FiO2 ratio | Respiratory rate, /min | Funding |
|---|---|---|---|---|---|---|---|
| three-arm, open-label, adaptive RCT, UK (75 centres)<br><br>N=1272 | treatment escalation was required | (primary comparisons were CPAP to standard oxygen and HFNO to standard oxygen) | during study period<br><br>Critical care (ICU) LOS<br><br>Hospital LOS | 57.6 ± 12.7 | 134.9 ± 82.8 | 25.0 ± 6.8 | National Institute for Health Research |
| **Teng et al. 2021**[39]<br><br>two-arm, parallel RCT, CHINA (single centre)<br><br>N= 22 | Patients diagnosed with severe COVID-19 | HFNO [n=12]<br><br>Standard oxygen therapy [n=10] | "Cured and discharged" (100% so used to infer not death)<br><br>Hospital LOS<br><br>ICU LOS | HFNC<br>56.6 ± 3.0<br><br>SOT<br>53.5 ± 5.5 | HFNC<br>224.25 ± 12.60<br><br>SOT<br>216.70 ± 4.62 | HFNC<br>22.08 ± 0.70<br><br>SOT<br>21.60 ± 0.40 | "The second batch of COVID-19 emergency science and technology project in Fuyang city (FK20202802)" |
| **Ospina-Tascón et al. 2021**[29]<br><br>Two-arm, open-label parallel RCT, Colombia, three centres<br><br>N=199 | Adult patients admitted to the emergency department, general ward, or intensive care unit with acute respiratory failure and COVID-19 | HFNO [n=99]<br><br>Standard oxygen therapy [n=100] | Mortality, 28 d<br><br>Intubation, 28 d<br><br>Hospital LOS<br><br>ICU LOS<br><br>No patient-reported outcomes | HFNO<br>60 (50-69)[a]<br><br>SOT<br>59 (49-67) | HFNO<br>104 (85-132)[a]<br><br>SOT<br>105 (85-141) | HFNO<br>28 (27-32)[a]<br><br>SOT<br>28 (26-31) | "The study received funds from the Centro de investigaciones Clínecas, Fundación Valle del Lili, Cali, Colombia." |

a: study reports median and interquartile range.

**Online supplement 5: Detailed RoB COVID RCTs**

*Figure S2. Risk of bias assessments for included COVID-19 RCTs.*

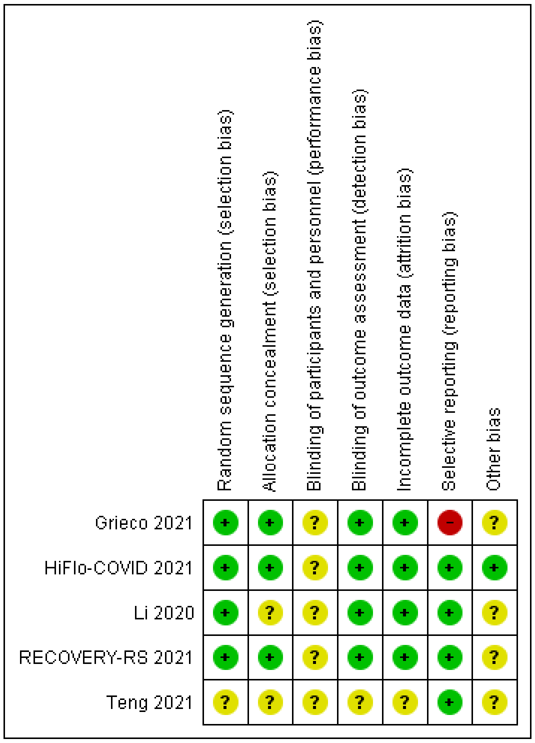


**Detailed RCT ROB assessments**

*Table S13. Risk of bias summary for Teng et al. 2020*

| Domain/ Description | Quote supporting judgement | Judgement |
|---|---|---|
| **Random sequence generation** | "Of these patients, 12 were randomized assigned to the HFNC oxygen therapy group and 10 were randomized assigned to the conventional oxygen therapy (COT) group…".<br><br>Methods for **sequence generation not described.** | Unclear |
| **Allocation concealment** | As above, method for a**llocation concealment not described.** | Unclear |
| Blinding of participants and personnel | Blinding not reported and likely impossible due to the use of different apparatus/techniques. The participants' and personnel's performance could have been biased due to their knowledge of the assigned treatment. | Unclear |
| Blinding of outcome assessors | Blinding not reported. | Unclear |

| | | |
|---|---|---|
| Incomplete outcome data - mortality | -- | Unclear |
| Incomplete outcome data-IMV | -- | Unclear |
| Selective outcome reporting | No protocol was not found. However, the outcomes of interest are reported as planned in the methods section. | Low |

*Table S14. Risk of bias summary for Grieco et al. 2021*

| Domain/ Description | Quote supporting judgement | Judgement |
|---|---|---|
| **Random sequence generation** | **"A computer-generated randomization scheme with randomly selected block sizes ranging from 3 to 9 managed by a centralized web-based system was used to allocate participants to each group."[3]** | **Low** |
| **Allocation concealment** | **As above, a centralized web-based system was used. It was judged appropriate.** | **Low** |
| Blinding of participants and personnel | **"...an investigator-initiated, 2-group, open-label, multicenter, randomized clinical trial..."**<br>No blinding. The participants' and personnel's performance could have been biased due to their knowledge of the assigned treatment. | **Unclear** |
| Blinding of outcome assessors | **"...Because the final decision on intubation was left to the physician in charge who could not be blinded to the study group, 2 independent experts blindly reviewed a posteriori the records and verified whether the decision to intubate was unbiased and in compliance with the required criteria. In case of disagreement between experts, a third physician established whether the criteria had been met."**<br>**No blinding but the intubation intervention followed strict and objective criteria and retrospectively reviewed and verified in consensus.** | **Unclear (IMV not adjudicated, not used)**<br>**Low (IMV adjudicated, mortality)**<br>**Both outcomes were presented** |
| Incomplete outcome data - mortality | **"...intensive care unit mortality, in-hospital mortality, 28-day mortality, 60-day mortality...Ninety-day mortality and quality of life after 6 and 12 months were** | **Low** |

[3] Some concern over baseline imbalance in Table 1.

| | | |
|---|---|---|
| | **among the pre-specified secondary outcomes, but results are not reported."**<br><br>**Mortality-related outcome assessment not likely influenced at all.** | |
| Incomplete outcome data-IMV | **As presented in figure 2, all randomized participants were included in the analysis except for two in the noninvasive ventilation helmet group and one in the high-flow nasal oxygen group, with the overall completion rate of 97% (107/110). It was judged to be at low risk of bias for incomplete outcome data.** | **Low** |
| Selective outcome reporting | **Protocol was registered (NCT02107183). However, the reported primary outcome, the number of days free of respiratory support (including high-flow nasal oxygen, noninvasive and invasive ventilation) within 28 days after enrollment", was different from what was pre-planned "Reintubation within 72 hours after extubation or at ICU discharge". Some of the secondary outcomes in the main publication were not described in the registered protocol, e.g., the number of days free of invasive mechanical ventilation at days 28 and 60. It was judged to be at high risk of reporting bias. Prespecified outcomes 90 mortality and quality of life not reported and no rationale provided.** | **High** |

*Table S15. Risk of bias summary for Li et al. 2020*

| Domain/ Description | Quote supporting judgement (copy from article with quotation marks) | Judgement |
|---|---|---|
| **Random sequence generation** | "７２例新型冠状病毒肺炎并发急性呼吸衰竭患者，按随机数字表(random number table)法将患者分为观察组与对照组。"<br>Random number table was used and judged to be appropriate. | Low |
| **Allocation concealment** | Method for **allocation concealment was not provided.** | Unclear |
| Blinding of participants and personnel | Blinding was not reported and appeared infeasible due to the two treatments involving different apparatus /techniques. The participants' and personnel's performance | Unclear |

| | | |
|---|---|---|
| | could have been biased due to their knowledge of the assigned treatment. | |
| Blinding of outcome assessors | The outcome "intubation after 12-hour continuous treatment" was investigated but the criteria were not provided. The personnel's administration/decision of intubation could have been based on participant's signs and symptoms and clinical judgement. | Low |
| Incomplete outcome data - mortality | No mortality outcomes reported. | Low |
| Incomplete outcome data-IMV | It appeared that all randomized participants were followed to the end of the study. No attrition was reported. | Low |
| Selective outcome reporting | Protocol was not available. However, the reported outcomes appeared to match the methods section. | Low |

*Table S16. Risk of bias summary for Perkins et al. 2021 (RECOVERY-RS)*

| **Domain/ Description** | **Quote supporting judgement (copy from article with quotation marks)** | **Judgement** |
|---|---|---|
| **Random sequence generation** | "Eligible participants were randomized using an internet-based system with allocation concealment…Randomization was stratified by site, sex, and age, and the allocation was generated by a minimization algorithm."<br>The method for sequence generation was judged appropriate. | Low |
| **Allocation concealment** | As above, allocation concealment was confirmed. | Low |
| Blinding of participants and personnel | "In this open-label, three-arm, adaptive, randomized controlled trial…"<br>No blinding. The participants' and personnel's performance could have been biased due to their knowledge of the assigned treatment. | Unclear |
| Blinding of outcome assessors | "Tracheal intubation was performed when clinically indicated, based on the judgement of the treating clinician."<br>Although blinding was not conducted, the administration/decision of intubation was based on participant's signs and symptoms and clinical judgement. | Low |
| Incomplete outcome data - mortality | "The primary outcome was a composite outcome of tracheal intubation or mortality within 30-days of randomization… The primary and secondary analyses were | Low |

| | | |
|---|---|---|
| | performed for the intention-to-treat (ITT) population…Primary outcome data were available for 99.0 % (1259/1272) of participants.”<br>The attrition was trivial, which was less likely to significantly influence the estimate of the effect size. | |
| Incomplete outcome data-IMV | As above, the attrition was trivial and outcome, intubation, was less likely to significantly influence the estimate of the effect size | Low |
| Selective outcome reporting | Trial protocol was posted online (statistical_analysis_plan_of_the_recovery-rs_trial_formal_v1.0_clean.pdf (warwick.ac.uk).The reported outcomes in the publication appear to match what have been pre-planned in the statistical analysis plan. | Low |

*Table S17. Risk of bias summary for Ospina-Tascón et al. 2021 (HiFLo-COVID)*

| **Domain/ Description** | **Quote supporting judgement (copy from article with quotation marks)** | **Judgement** |
|---|---|---|
| Random sequence generation | "Randomization was centrally performed through a web-based system using computer-generated random numbers with blocks of 2 and 4, unknown to the investigators, and was stratified by study site to ensure allocation concealment." | Low |
| Allocation concealment | "Randomization was centrally performed through a web-based system using computer-generated random numbers with blocks of 2 and 4, unknown to the investigators, and was stratified by study site to ensure allocation concealment." | Low |
| Blinding of participants and personnel | "Participating patients could not be masked because of the nature of the intervention." | Unclear |
| Blinding of outcome assessors | "This study has several limitations. First, because of its nature, this open-label trial lacked the possibility of blinding, which may affect the assessment of outcomes."<br><br>"Nevertheless, main investigators were unaware of the study group outcomes until the database was locked after the end of follow-up on February 10, 2021. An independent statistician performed all the analyses." | Low |

| | | |
|---|---|---|
| Incomplete outcome data - mortality | No concerns | Low |
| Incomplete outcome data-IMV | No concerns | Low |
| Selective outcome reporting | No concerns | Low |

**Online supplement 6: Detailed outcome tables**

***Mortality***

*Table S18. MORTALITY - HFNO versus STANDARD OXYGEN*

| STUDY | HFNO | | SOT | |
|---|---|---|---|---|
| | n | N | n | N |
| HiFLo COVID | 8 | 99 | 16 | 100 |
| Teng 2021 | 0 | 12 | 0 | 10 |
| RECOVERY-RS | 78 | 415 | 74 | 370 |

*Table S19. MORTALITY - CPAP versus STANDARD OXYGEN*

| STUDY | CPAP | | SOT | |
|---|---|---|---|---|
| | n | N | n | N |
| RECOVERY-RS | 63 | 378 | 69 | 359 |

*Table S20. MORTALITY - HELMET NIV versus HFNO*

| STUDY | HELMET NIV | | HFNO | |
|---|---|---|---|---|
| | n | N | n | N |
| GRIECO 2021 | 8 | 55 | 10 | 55 |

*Table S21. MORTALITY - CPAP versus HFNO*

| STUDY | CPAP | | HFNO | |
|---|---|---|---|---|
| | n | N | n | N |
| RECOVERY-RS | 63 | 378 | 69 | 359 |

***IMV***

*Table S22. IMV - HFNO versus STANDARD OXYGEN*

| STUDY | HFNO | | SOT | |
|---|---|---|---|---|
| | n | N | n | N |
| HiFLo COVID | 34 | 99 | 51 | 100 |
| Li 2020 | 1 | 37 | 6 | 35 |
| RECOVERY-RS | 170 | 414 | 153 | 368 |

*Table S23. IMV - CPAP versus STANDARD OXYGEN*

| STUDY | CPAP | | SOT | |
|---|---|---|---|---|
| | n | N | n | N |
| RECOVERY-RS | 126 | 377 | 147 | 356 |

*Table S24. IMV - HELMET NIV versus HFNO*

| STUDY | HELMET NIV | | HFNO | |
|---|---|---|---|---|
| | n | N | n | N |
| GRIECO 2021 | 15 | 55 | 28 | 55 |

*Table S25. IMV - CPAP versus HFNO*

| STUDY | CPAP | | HFNO | |
|---|---|---|---|---|
| | n | N | n | N |
| RECOVERY-RS | 126 | 377 | 170 | 414 |

***Hospital and ICU LOS***

*Table S26. HOSPITAL AND ICU LOS - HFNO versus STANDARD OXYGEN*

| HOSPITAL LOS | | | | | | |
|---|---|---|---|---|---|---|
| STUDY | HFNO | | | SOT | | |
| | days, mean | standard deviation | N | days, mean | standard deviation | N |
| HiFLo COVID | 12[a] | 9-20[b] | 99 | 14[a] | 9-23[b] | 100 |
| Teng 2021 | 14.67 | 1.97 | 12 | 16.6 | 2.54 | 10 |
| RECOVERY-RS | 18.3 | 20 | 414 | 17.1 | 18 | 368 |
| ICU LOS | | | | | | |
| STUDY | HFNO | | | SOT | | |
| | days | variation | N | days | variation | N |
| HiFLo COVID | 7[a] | 5-13[b] | 99 | 9[a] | 5-18[b] | 100 |
| Teng 2021 | 4 | 0.74 | 12 | 4.9 | 1 | 10 |
| RECOVERY-RS | 10.5 | 15.6 | 414 | 9.5 | 14.1 | 368 |

a:media; b:interquartile range

*Table S27. HOSPITAL AND ICU LOS - CPAP versus STANDARD OXYGEN*

| HOSPITAL LOS | | | | |
|---|---|---|---|---|
| STUDY | CPAP | | SOT | |
| | days, mean | standard deviation | days, mean | standard deviation |
| RECOVERY-RS | 16.4 | 17.5 | 17.3 | 18.1 |
| ICU LOS | | | | |
| STUDY | CPAP | | SOT | |
| | days, mean | standard deviation | days, mean | standard deviation |
| RECOVERY-RS | 9.5 | 15.6 | 9.6 | 13.6 |

*Table S28. HOSPITAL AND ICU LOS - HELMET NIV versus HFNO*

| HOSPITAL LOS | | |
|---|---|---|
| STUDY | HELMET NIV | HFNO |

| | days, median | IQR | days, median | IQR |
|---|---|---|---|---|
| GRIECO 2021 | 21 | 14-30 | 22 | 13-44 |
| **ICU LOS** | | | | |
| **STUDY** | **HELMET NIV** | | **HFNO** | |
| | **days, median** | **IQR** | **days, median** | **IQR** |
| GRIECO 2021 | 9 | 4-17 | 10 | 5-23 |

*Table S29. HOSPITAL AND ICU LOS - CPAP versus HFNO*

| **HOSPITAL LOS** | | | | |
|---|---|---|---|---|
| **STUDY** | **CPAP** | | **HFNO** | |
| | **days, mean** | **SD** | **days, mean** | **SD** |
| RECOVERY-RS | 16.4 | 17.5 | 18.3 | 20 |
| **ICU LOS** | | | | |
| **STUDY** | **CPAP** | | **HFNO** | |
| | **days, mean** | **SD** | **days, mean** | **SD** |
| RECOVERY-RS | 9.5 | 15.6 | 10.5 | 15.6 |

**Meta-analysis: Tables**

***Indirect data calculations***

*Table S30. Indirect data calculations for CPAP vs HFNO exploratory analyses (Specific to the RECOVERY-RS RCT)*

| **OUTCOME** | **RCT DATA REPORTED** | | **INDIRECT TREATMENT COMPARISON** |
|---|---|---|---|
| | **HFNO versus SOT** | **CPAP versus SOT** | **CPAP versus HFNO (ITC DATA)** |
| **MORTALITY AT 30 D** | ADJUSTED OR = 0.96 (0.64 - 1.45) | ADJUSTED OR = 0.91 (0.59 -1.39) | RR 0.948 (0.524-1.714) |
| **IMV AT 30D** | ADJUSTED OR 0.96 (0.70 - 1.31) | ADJUSTED OR = 0.66 (0.47- 0.93) | RR 0.688 (0.433-1.093) |
| **HOSPITAL LOS, D** | MD 0.70 (-1.93, 3.34) | MD -0.97 (-3.65, 1.71) | MD -1.67 (-5.428, 2.088) |
| **ICU LOS, D** | MD 0.69 (-1.37, 2.75) | MD -0.33 (-2.44, 1.78) | MD -1.02 (-3.969, 1.929) |

***Results for length of stay outcomes***

*Table S31. META-MEDIAN OUTPUT: Pooled data by arm from meta-analysis - Hospital and ICU LOS: HFNO versus SOT (fixed effects)*

| OUTCOME | NO. RCTS | HFNO | | | | | | SOT | | | | | |
|---|---|---|---|---|---|---|---|---|---|---|---|---|---|
| | | DAYS, MEAN | LCI | UCI | SE | N | $I^2$ | DAYS, MEAN | LCI | UCI | SE | N | $I^2$ |
| HOSPITAL LOS, D | 3 | **14.92** | 14.04 | 15.79 | 0.44 | 525 | 90% | **16.28** | 15.20 | 17.36 | 0.55 | 478 | 52% |
| ICU LOS, D | 3 | **4.65** | 4.26 | 5.04 | 0.20 | 525 | 97% | **5.83** | 5.27 | 6.38 | 0.28 | 478 | 95% |

*Table S32. META-MEDIAN OUTPUT: Meta-analysis - Hospital and ICU LOS: HFNO versus SOT (fixed effects), absolute mean difference, in days*

| OUTCOME | NO. RCTS | ABSOLUTE MEAN DIFFERENCE, D | LCI | UCI | SE | N | $I^2$ |
|---|---|---|---|---|---|---|---|
| HOSPITAL LOS, D | 3 | **-1.08** | -2.48 | 0.33 | 0.72 | 1003 | 48% |
| ICU LOS, D | 3 | **-0.77** | -1.45 | -0.09 | 0.35 | 1003 | 46% |
